# Determination of significant immunological timescales from mRNA-LNP-based vaccines in humans

**DOI:** 10.1101/2022.07.25.22278031

**Authors:** Iain R. Moyles, Chapin S. Korosec, Jane M. Heffernan

**Author notes:** Contributing authors.

## Abstract

A compartment model for an in-host liquid nanoparticle delivered mRNA vaccine is presented. Through non-dimensionalisation, five timescales are identified that dictate the lifetime of the vaccine in-host: decay of interferon gamma, antibody priming, autocatalytic growth, antibody peak and decay, and interleukin cessation. Through asymptotic analysis we are able to obtain semi-analytical solutions in each of the time regimes which allows us to predict maximal concentrations and better understand parameter dependence in the model. We compare our model to 22 data sets for the BNT162b2 and mRNA-1273 mRNA vaccines demonstrating good agreement. Using our analysis, we estimate the values for each of the five timescales in each data set and predict maximal concentrations of plasma B-cells, antibody, and interleukin. Through our comparison, we do not observe any discernible differences between vaccine candidates and sex. However, we do identify an age dependence, specifically that vaccine activation takes longer and that peak antibody occurs sooner in patients aged 55 and greater.

## 1 Introduction

Vaccines are one of the greatest advents of modern society, significantly reducing mortality (cf. Rodrigues and Plotkin (2020)) with Ehreth (2003) estimating a prevention of 6 million deaths of vaccine-preventable diseases each year. Vaccines are derived from many precursors including from inactivated virus, viral protein subunits, recombinant human adenovirus, and messenger RNA (mRNA) with the latter being of increasing interest due to their high potency, low manufacturing costs, and the ability to be developed quickly as outlined by Pardi et al (2018b). mRNA vaccines gained prominence during the COVID-19 pandemic with the development and deployment of BNT162b2 (produced by Pfizer-BioNTech) and mRNA-1273 (produced by Moderna) both of which are injected via liquid nanoparticles (LNP). This mechanism was chosen to aid in cell delivery and protect the mRNA from degradation (cf. Ndeupen et al (2021)).

mRNA vaccines have been in development for many years. A review of their usage in infectious diseases has been conducted by Zhang et al (2019). Prior to COVID-19 it was recognized that mRNA vaccines were outperforming other technologies such as inactivated virus and protein adjuvanted vaccines (cf. Pardi et al (2018a)). mRNA vaccines have also demonstrated robust immune responses to other diseases. A study by Bahl et al (2017) showed vaccines to be effective against severe disease of H7N9 and H10N8 influenza viruses in mice, non-human primates, and ferrets.

mRNA vaccines, as with other vaccine types, demonstrate waning effectiveness. A study by Menni et al (2022) concluded that antibody protection from COVID-19 mRNA vaccines showed significant waning beginning 5 months after the standard two dose regiment, acknowledging that overall vaccine effectiveness was dependent on age and comorbidity. However, they also saw continued protection from severe disease beyond 6 months. Clinical studies such as these demonstrate the importance of measuring immunity response, development, and decay in mRNA vaccines for COVID-19. However, these trials can be very costly and, without a clear understanding of the immune response, the data collection requirements can be uncertain. Mathematical modelling and analysis provides a cost-effective tool for understanding and predicting the immunity response to vaccines.

Most mathematical modelling of infectious disease occurs at the population scale and throughout the COVID-19 pandemic there have been many such studies (cf. Tang et al (2020); Moyles et al (2021); Yuan et al (2022a); Fair et al (2022); Childs et al (2022); Vignals et al (2021); Betti et al (2021); Dick et al (2021); Moore et al (2021); Moss et al (2020); Smirnova et al (2021); Wells et al (2021); Li et al (2020); Yuan et al (2022b); Hogan et al (2021)). While the impacts of vaccine efficacy and waning are important at these scales, the actual process of immune development occurs within-host. In-host mathematical modelling considers pathogen reproduction and cellular infection within a single individual and has been effectively employed in various diseases. For example, Heffernan and Keeling (2009) modelled vaccination and waning with measles, Herz et al (1996) modelled the intracellular viral life cycle phase of HIV and hepatitis B, and Perelson (2002) reviewed immune system dynamic modelling for HIV, hepatitis C, and cytomegalovirus (CMV). In-host modelling has been extended to COVID-19 with studies looking at infection (cf. Hernandez-Vargas and Velasco-Hernandez (2020); Perelson and Ke (2021); Kim et al (2021); Néant et al (2021); Sadria and Layton (2021); Lin et al (2022); Ke et al (2022); Korosec et al (2023)) and vaccination (cf. Farhang-Sardroodi et al (2021); Korosec et al (2022); Gholami et al (2023)).

In this paper we explore an in-host mathematical model for an LNP vaccine first considered by Korosec et al (2022) where computational mixed-effects modelling was used to identify model parameters from a variety of data sets. Their results demonstrated a diverse variability in the parameter estimates and consequently the model comparisons. Furthermore, the nature of the clinical trial data analyzed meant that the time series had coarse resolution spread over a long duration. In this paper, we identify the dominant terms of the model and the timescales over which they are relevant with the goal of elucidating the antibody process and providing direction for improved data resolution. However, we also provide mechanistic insight into the immune response which is important because the precise timescales of activating the immune response in humans is relatively unknown. Luceripherase mouse studies have been conducted, for example, by Pardi et al (2015) and Lutz et al (2017) showing that the injection site remains active for approximately 7-10 days and that luceriphase was also present in the draining lymph nodes nearest the injection site a few days after injection. Similar luceriphase studies cannot be done on humans and instead highly resolved blood draws are needed to monitor the adaptive and innate immune response. Our timescale analysis provides a framework to understand these physiological processes sequentially.

Understanding a sequential ordering of processes that drive vaccine dynamics is very beneficial. It helps understand and separate the innate immune responses from the adaptive ones generated by the vaccine. Separating these timescales helps identify balance between the immune responses which is important for limiting the body’s ability to attack vaccine species at dosage. A thorough model analysis also improves mechanistic understanding. Even though mRNA vaccines have demonstrated strong immune responses, the immunological activity leading to these responses is less clear (cf. Lindgren et al (2017)). Our analysis generalizes beyond mRNA vaccines as well. The model we present is itself adapted from an adenovirus-based vaccine. Therefore, our work can be used to broadly decompose vaccine-mediated immunity for a variety of candidates and diseases.

A common approach in immunological modelling studies is to focus solely on simulations. Our approach exploits asymptotic methods to isolate model structure and function as it evolves with time identifying important parameter relationships. While this methodology is mathematically richer than solely a simulation based study, it can also provide practical and biological insight. For example, parameter identifiability is an important consideration in modelling studies. Analytic expressions determine explicit relationships with parameters which can elucidate both model identifiability, i.e., which parameters form scaling groups, as well as practical identifiability, i.e., what time points are needed in data collection to identify certain parameters. Formal asymptotic methods can justify the inclusions or removal of certain terms in a model adding rigor and confidence to biological intuition. Analytic methods can enrich biological understanding of a problem by linking processes with explicit parameter relationships. For example, understanding a peak immunological response explicitly in terms of model parameters can provide insight into the processes that drive this response while also allowing researchers to exploit new vaccines and therapies to improve the response.

The paper is organized as follows. We introduce and summarize the model of Korosec et al (2022) in Section 2. We nondimensionalize the model in Section 2.1 where, through some assumption of scales, we are able to reduce the model to a simpler form. We identify a series of timescales in Section 3 that account for antibody development, growth, maximum, and ultimately decay. In each timescale regime we are able to determine asymptotic analytic solutions that explain the immunity response. We compare these analytic results to a full numerical simulation of the model in Section 4 where maxima of key immunity factors are approximated and we compare the model to data sets from a series of clinical trials. We discuss results and conclude the paper in Section 5.

## 2 Model Summary and Reduction

We consider a compartmental ordinary differential equation model first published by Farhang-Sardroodi et al (2021) for adenovirus-based vaccines and later adapted by Korosec et al (2022) for LNP mRNA vaccines. The process being modelled begins with the injection of a concentration of LNP (*L*) that diffuses through inactive target cells of which we assume there is an infinite reservoir. Details of these target cells are unclear. When injected intramuscularly, such as with humans, Lutz et al (2017) found in a study of mice that mRNA vaccines activated the innate immune system near the site of injection as well as in draining lymph nodes. These target cells activate to become vaccinated cells (*V*) that promote the production of CD4^+^ (*T*) and cytotoxic CD8^+^ (*C*) T-cells. These CD4^+^ T-cells further promote the production of plasma B-cells (*B*), interferon-*γ* (IFN-*γ, F*), and interleukin (*I*). Finally, the plasma B-cells stimulate production of immunoglobulin G (IgG) antibody (*A*). The equations of the model studied by Korosec et al (2022) are given by

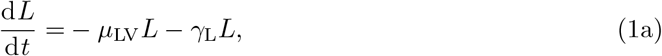

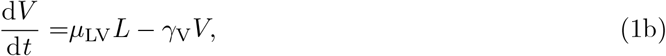

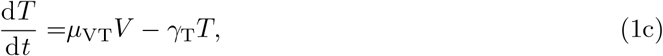

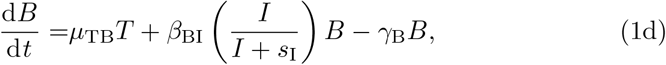

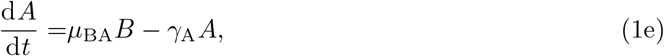

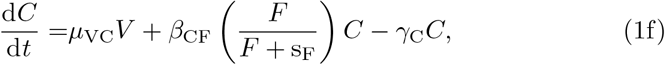

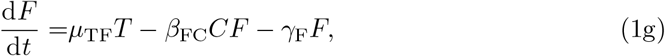

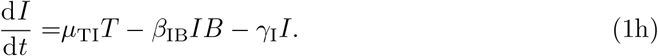

The parameters *μ*_*ij*_ are priming rates (d^−1^) that activate component *j* from component *i. β*_*ij*_ are immune response autocatalytic rates (d^−1^) arising from non-linear activation and inhibition of species *j* on species *i*, and *γ*_*i*_ are the natural decay rates (d^−1^). The parameters *s*_I_ and *s*_F_ are concentrations of interleukin and IFN-*γ*, respectively, that produce half-maximal autocatalytic rates, *β*_*ij*_. Time, *t*, is measured in days and concentrations of each component are in arbitrary volumetric units. We note that equations (1d) and (1f) contain saturation dynamics through the terms *s*_*F*_ and *s*_*I*_. We include these to be consistent with the full model of Korosec et al (2022), but biologically it will be important to limit activating kinetics in a general model of vaccination which could include multiple doses.

The only feasible steady state to (1) is every concentration being zero and thus, in the absence of some initial vaccine, it is reasonable within the context of the model to measure no immune response. However, it is possible through previous infection or other biological mechanisms not accounted for in the model, that some basal concentrations may exist for quantities not primed through vaccine. As such, we consider the generalized initial conditions to (1) to be

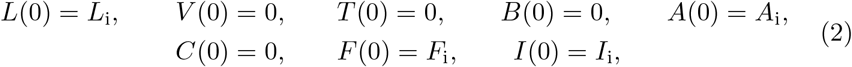

where we have assumed that vaccine is injected at time *t* = 0 with concentration *L*_i_.

### 2.1 Non-dimensionalization and model reduction

To understand the effects of each term in the model we non-dimensionalize (1). Firstly, the concentration of LNP will be determined by its initial value as it only decays and thus we scale *L* ∼ *L*_i_. Secondly, as the dynamics are driven by vaccination, the natural time scale is the activation of vaccinated cells by LNP and thus we scale 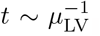. We do not scale other variables by their initial condition because we expect significant variability in these levels between individuals. Furthermore, since the model is derived based on vaccine response, we anticipate that it should be driven by the dynamics of vaccination rather than any latent initial concentrations present. Instead, we look to (1) and choose scales by balancing source and sink terms in the model. For example in (1b), (1c), (1d), (1f), and (1h) vaccine priming drives production and so it is sensible to choose scales for *V, T, B, C*, and *I* driven by the source terms. For IgG in (1e) it is sensible to balance the source of plasma cells with decay of antibody, *A*. For *F* in (1g), we scale the concentration of IFN-*γ* assuming its natural decay is the dominant sink term. This is consistent with fast decay rates of IFN-*γ* of between 3 and 40 minutes in mice and humans reported by Foon et al (1985); Gonias et al (1988); Psimadas et al (2012). We note that this, and all decay terms, strictly refers to the removal from the vaccine induced antibody response being modelled here. It does not necessarily mean that the components are degrading, but could instead by absorbed or consumed in other biological functions not related to immune response of the vaccine. Overall, we are led to the following scales for each of the concentrations

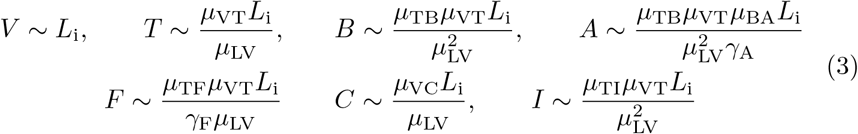

which leads to the non-dimensional model

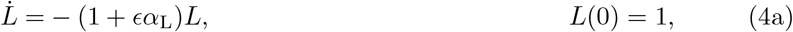

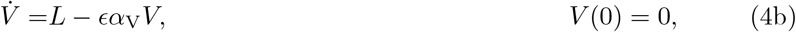

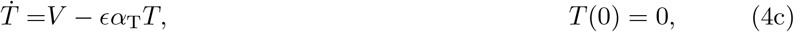

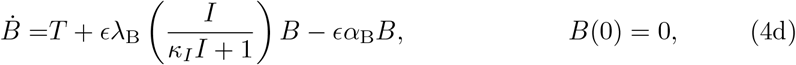

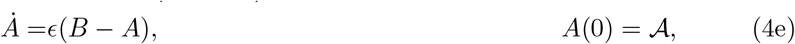

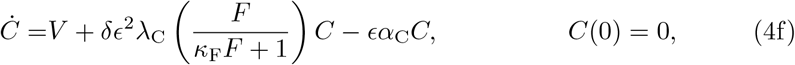

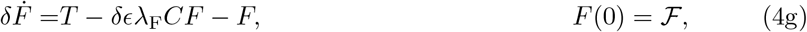

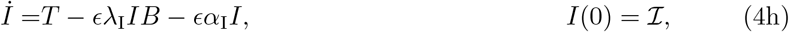

where dot indicates differentiation with respect to non-dimensional time. The parameters in (4) are defined as follows:

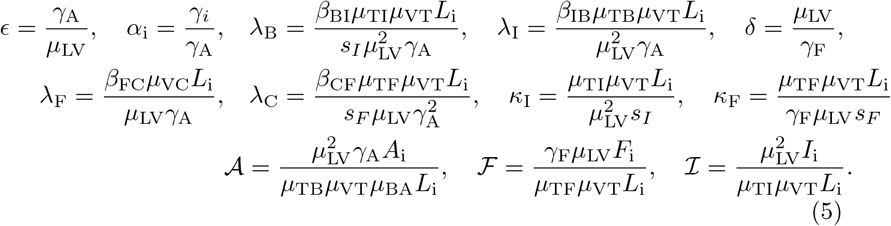

Many of these parameters have natural interpretations. For example *ϵ* is the timescale ratio of vaccine absorption to decay of IgG, the terminal antibody; the parameter *α*_*i*_ is the timescale ratio of the natural decay of concentration *i* relative to the decay of IgG; and the parameter *δ* is the timescale ratio of release of vaccine to decay of IFN-*γ*. The parameters *κ*_I_ and *κ*_F_ are effective saturation constants which limit the production of plasma B cells and cytotoxic T-cells while the parameters *λ*_*i*_ are the strengths of the autocatalytic production terms compared to the vaccine priming sources for the *B, C, F*, and *I* compartments. Each of 𝒜, ℱ, and ℐ are non-dimensional initial concentrations of IgG, IFN-*γ*, and interleukin respectively.

We anticipate that an effective immune response necessitates that the decay of antibodies is much slower than the absorption of vaccine in the LNP and thus that *ϵ* ≪ 1. This is supported experimentally where Lutz et al (2017), for example, observed that mice began to produce antigen at the site of injection on the order of hours after the needle, while antibody decay halflife was on the order of many days. Conversely, because of the fast decay rates on the order of minutes of IFN−*γ* observed by Foon et al (1985) and others compared to the LNP absorption timescale of hours observed by Lutz et al (2017), we will assume that *δ* ≪ 1 as well. We anticipate that the ratio of the remaining decay rates to *γ*_*A*_ are comparable and thus take *α*_*i*_ ∼ 𝒪(1). Without any presupposition about the magnitude of production multipliers, *λ*_*i*_, we simply assume formally that they are 𝒪(1). So long as terms are not an order of magnitude larger than we have assumed, the model will be consistently scaled and its structure and analysis will be valid. The actual size of terms is dependent on specific case studies, a detail we resolve in Section 4.2.

Assuming the saturation parameters *κ*_*i*_ are small is generally associated with limited saturation effect. While this is likely true in (4g) where IFN-*γ* decays quickly, it is less clear in (4d) where autocatalytic production of interleukin may significantly contribute to antibody production. However, if saturation effects become important then this is likely a consequence of an overactive immune system and we assume that is generally not the case.

Based on the assumptions, we propose a reduced model to (4) where *κ*_I_ = *κ*_F_ = 0. Furthermore, since both *ϵ* ≪ 1 and *δ* ≪ 1 then the terms associated to *λ*_C_ in (4f) and *λ*_F_ in (4g) are likely non-identifiable and therefore, we will assume that *λ*_F_ = *λ*_C_ = 0 without loss of generality. Finally in (4a) since we have two competing sink terms we assume the second, being the smaller of the two, is non-identifiable. Thus we will assume that *α*_L_ = 0. Overall, we are led to the following reduced model:

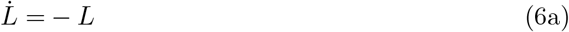

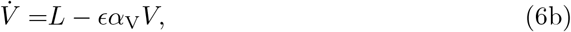

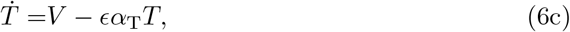

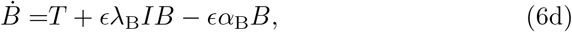

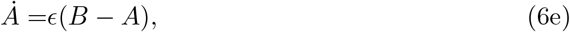

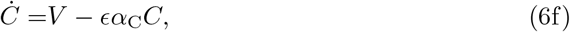

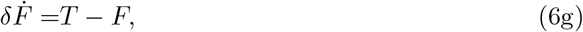

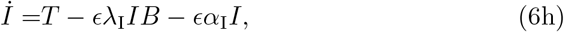

subject to the same initial conditions as (4). It may seem unsatisfactory that we have neglected some small terms in the reduced model (6) while leaving others. This is justified from (6e) where if we take *ϵ* ≡ 0 there is no mechanism for antibody response. Therefore, necessarily, there must be some long timescale antibody production driven by *ϵ*. Thus, we leave the small terms associated to this parameter. We note that this is also consistent with not scaling terms by latent initial conditions. We demonstrate agreement between the full model (4) and the reduced model (6) in Section 4.

## 3 Timescale Decomposition

We will now analyze the reduced model (6) to understand the impact of different parameters on the long-term antibody response. We begin by noting that *L, V, T*, and *C* in (6) can be solved analytically as these are a cascade of linear equations that decouple from one another. The solutions, subject to the appropriate initial conditions *L*(0) = 1 and *V* (0) = *T* (0) = *C*(0), are

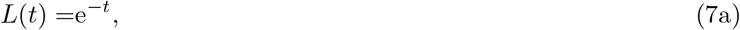

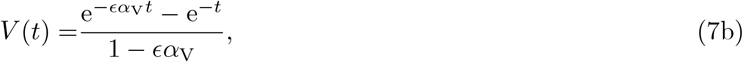

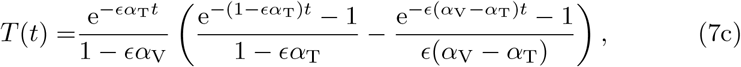

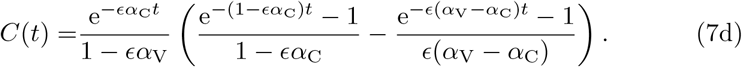

We note in model (7) we have made the explicit assumption that *α*_T_ ≠ *α*_V_ and *α*_C_ ≠ *α*_V_ for generality. We will use the assumption that all *α*_V_, *α*_T_, and *α*_B_ are unique throughout the remainder of the manuscript and discuss the specific case where the parameters are equal in Appendix B.

The anti-body concentration, (6e) depends on the plasma B-cell concentration, *B*, and thus can not yet be solved in closed form. However, we can write its solution in terms of the plasma B-cell concentration,

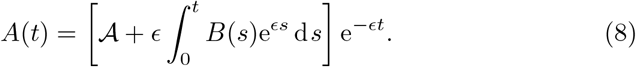

We note that we can also solve (6g) which has the general solution,

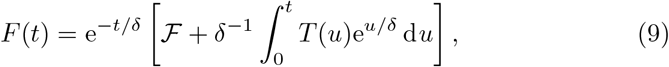

with *T* given by (7c). However, aside from a fast timescale of 𝒪(*δ*) as described in section 3.1, this equation is mostly in quasi-steady state with

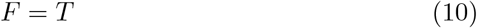

which is more insightful than the full solution (9).

The solutions (7), (8), and (9) already significantly decouple the eight equation model (6) to a two equation model for *B*, and *I* which we now systematically explore through a series of chronological time scales. The full description of these timescales is presented and derived in sections 3.1 to 3.5, but are also summarized in Table 1 for brevity.

**Table 1:** Summary of chronological timescales discussed in Sections 3.1 to 3.5.

| Timescale | Description |
| --- | --- |
| $t = \delta t_1$ (sec 3.1) | **Clearance of IFN-γ:** The IFN-γ is quickly cleared from the vaccine-induced immune response. This timescale removes any latent IFN-γ that exists as an initial condition. If $\mathcal{F} = 0$ then this timescale is irrelevant. |
| $t$ (sec 3.2) | **Vaccine Priming:** This is the timescale chosen to be $\mathcal{O}(1)$ in the non-dimensionalisation and therefore represents that absorption time of the vaccine. During this time, early production of interleukin and plasma B-cells is primed by the vaccine. To leading order there is no antibody production. |
| $t = \epsilon^{-1/3} t_3$ (sec 3.3) | **Antibody autocatalytic growth:** During the priming phase, vaccine induced production of plasma B-cells and interleukin activates the body's natural production mechanism leading to explosive growth of each constituent and the production of significant antibody. |
| $t = \epsilon^{-1} t_4$ (sec 3.4) | **Peak and Decay of Antibody:** The inhibiting effect of interleukin on its self-production leads to terminal production of plasma B-cells and thus terminal production of antibody. During this timescale all concentration production slows and decay becomes dominant. This mechanistic transfer leads to the peak antibody response occurring in this timescale. Depending on the decay kinetics of plasma B-cells and CD4<sup>+</sup> T-cells, a balancing between production and removal of interleukin occurs leading to a quasi-steady state interleukin concentration. |
| $t = \epsilon^{-1} \left( -\frac{2}{\alpha_I} \log \epsilon + t_5 \right)$ (sec 3.5) | **Interleukin cessation:** The model necessitates that all concentrations decay to zero in the absence of continued vaccine supply. This timescale resolves the quasi-steady state concentration of interleukin allowing it to decay to zero. |

### 3.1 IFN-*γ* Clearance

The first time scale emerges in (6g) when *t* ∼ 𝒪(*δ*), recalling that *δ* is the ratio of vaccine absorption to IFN-*γ* decay. As such we let *t* = *δt*_1_ resulting in every equation of (6) to effectively be in equilibrium except for (6g) which becomes

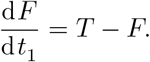

The CD4^+^ T-cells at this timescale are given by (7c) after substituting *t* = *δt*_1_ and expanding for *δ* ≪ 1. This results in *T* ≈ 0 and thus that

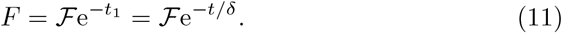

Therefore, at this timescale we have that any initial concentration of IFN-*γ* quickly clears the body. Conversely, if there is no initial concentration of IFN-*γ* then this timescale can be ignored as none of the other components have yet activated, since *B* and *I* are given by their initial conditions to leaing order when *t* ∼ 𝒪(*δ*).

### 3.2 Vaccine Priming

The next time scale occurs when *t* ∼ 𝒪(1), i.e. the selected timescale from the non-dimensionalisation. Having already removed terms via (7) and (8), and recalling that *F* = *T* from the quasi-steady limit (10), the reduced model at this scale is

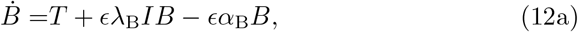

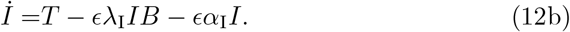

These equations are non-linear and do not have direct analytic solutions. Exploiting the smallness of the parameter *ϵ* we can expand (7c) when *t* ∼ 𝒪(1) and *ϵ* ≪ 1 to get,

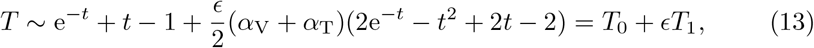

for the CD4^+^ T-cell population at this timescale. The formulation of this asymptotic series suggests we pose expansions *B* = *B*_0_ + *ϵB*_1_ and *I* = *I*_0_ + *ϵI*_1_ and substitute into (12). The leading order problem becomes

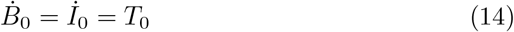

subject to *B*_0_(0) = 0 and *I*_0_(0) = ℐ which has solution

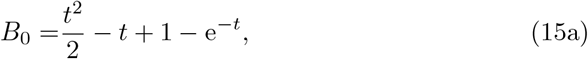

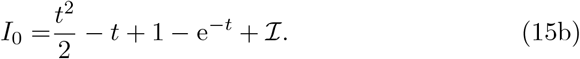

Expanding (12) to 𝒪(*ϵ*) we get

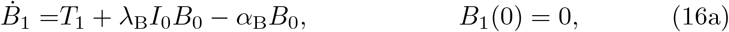

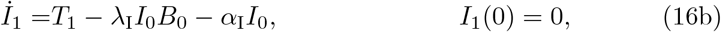

which has solution

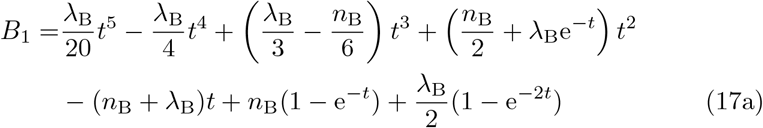

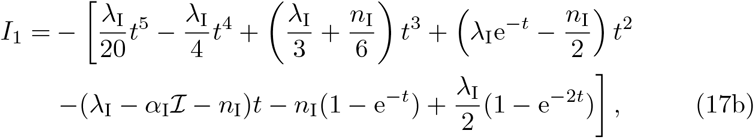

where we define

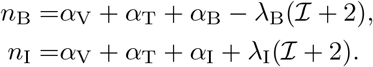

Having determined *B*, the anti-body concentration is given by expanding (8) for *t* ∼ 𝒪(1) and *ϵ* ≪ 1 which yields,

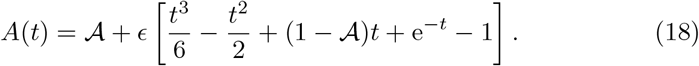

The leading order anti-body response is given by its initial condition and thus there is very little production at the *t* ∼ 𝒪(1) timescale. This result supports the physiological intuition that, at the 𝒪(1) timescale, the plasma B-cells and interleukin have yet to develop. Thus, antibodies are close to their background levels during this timescale.

The need for a longer time scale beyond *t* ∼ 𝒪(1) is immediately clear as each of the concentrations *T, B, A*, and *I* grow without bound. From (13), *T* loses asymptotic consistency when *t* ∼ 𝒪(*ϵ*^−1^) because the correction term grows quicker than the leading order term. This represents a time where the production of CD4^+^ T cells from the vaccine has stopped and only decay of remaining cells is taking place. However, from (17a) the plasma B-cell concentration loses asymptotic consistency when *t* ∼ 𝒪(*ϵ*^−1*/*3^), an earlier time than *t* ∼ 𝒪(*ϵ*^−1^). This breakdown is also evident in (17b) for interleukin and (18) for IgG antibody.

The early breakdown of the solution to the plasma B-cell concentration is due to the rapid autocatalytic production of plasma B-cells as interleukin concentration increases. Therefore, *t* ∼ 𝒪(*ϵ*^−1*/*3^) represents a switchover from the vaccine production of plasma B-cells being the dominant contribution to the self-stimulating autocatalytic production becoming dominant. We find that this breakdown happens beyond the leading order dynamics necessitating the two-term asymptotic expansion posed at this timescale.

### 3.3 Antibody Autocatalytic Growth

We introduce the new timescale *t* = *ϵ*^−1*/*3^*t*_3_, substitute into (7c) and take *ϵ* ≪ 1 to get that the T-cell concentration at this scale is,

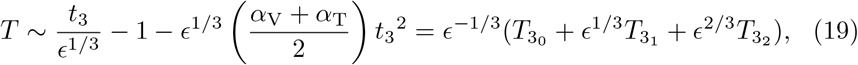

where we have scaled *T* = *ϵ*^−1*/*3^*T*_3_. Unsurprisingly, we see that *T*_3_ loses asymptotic consistency when *t*_3_ ∼ 𝒪(*ϵ*^−2*/*3^) equivalent to the asymptotic breakdown when *t* ∼ 𝒪(*ϵ*^−1^) as discussed in Section 3.2. Substituting the *t*_3_ timescale into (15b) and (15a) shows that it is sensible to scale *I* = *ϵ*^−2*/*3^*I*_3_ and *B* = *ϵ*^−2*/*3^*B*_3_. This leads to the reduced model (6) at this scale

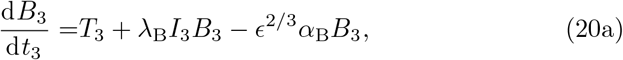

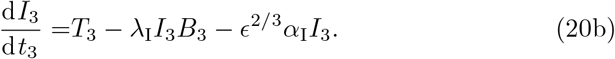

Unlike the 𝒪(1) time in Section 3.2, the leading order solution will be sufficient for capturing the dynamics at this timescale (see Appendix A) and this leading order is given by

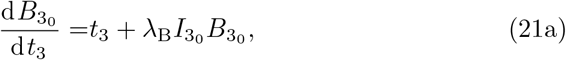

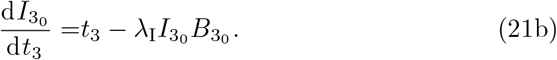

We obtain the leading order antibody concentration by substituting *t* = *ϵ*^−1*/*3^*t*_3_ into (8) and expanding for *ϵ* ≪ 1 yielding,

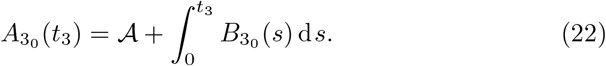

Multiplying (21a) by *λ*_I_ and (21b) by *λ*_B_ and adding yields

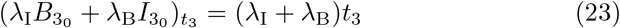

and thus

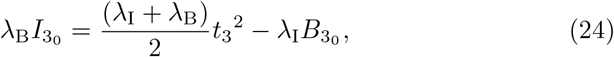

where we have chosen the integration constant so that both *B*_3_ and *I*_3_ tend to *t*_3_^2^*/*2 as required for matching to (15a) and (15b) when *t*_3_ ≪ 1. Substituting (24) into (21a) yields

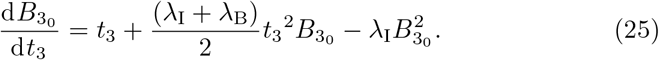

This non-linear equation can be solved using Kummer functions (see Appendix A) leading to the solution

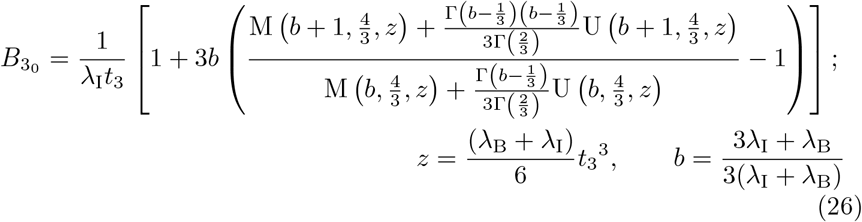

with M(*x, y, z*) and U(*x, y, z*) Kummer functions of the first and second kind respectively and Γ(*z*) the usual Gamma function (cf. Abramowitz and Stegun (1983); Polyanin and Zaitsev (2017)). The leading order solution (26) satisfies the correct matching condition that 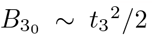 for *t*_3_ ≪ 1 (see (A9) in Appendix A for details) coming from the far-field behaviour of the solution (15a) in the *t* ∼ 𝒪(1) timescale. Having determined 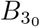 then 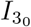 is determined from (24).

We show in Section A.1 of Appendix A that the solutions at this timescale do not lose asymptotic consistency prior to *t* ∼ 𝒪(*ϵ*^−1^) when the CD4^+^ T-cell concentration given by (19) loses asymptotic consistency and therefore there are no additional intermediate timescales for antibody growth to consider. This resolves the loss of asymptotic consistency for *B, I*, and *A* that was discovered at the *t* ∼ 𝒪(1) timescale in Section 3.2. However, the loss of asymptotic consistency in the CD4^+^ T-cell concentration is still unresolved as that occurred when *t* ∼ 𝒪(*ϵ*) due to the decay terms not being included.

### 3.4 Peak and Decay of Antibody

Introducing the timescale *t* = *ϵ*^−1^*t*_4_, we expand (7c) for *T* yielding,

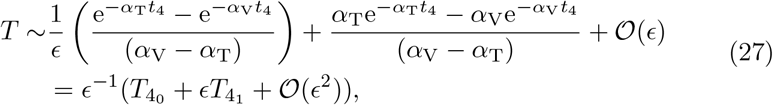

where we have scaled *T* = *ϵ*^−1^*T*_4_. In Appendix A we show the behaviour of *B, A*, and *I* when *t*_3_ ≫ 1 given by (A20), (A17) and (A21) respectively. This provides the matching scaling at the *t*_4_ timescale and therefore suggests we scale *B* = *ϵ*^−2^*B*_4_ and *I* = *I*_4_ transforming the reduced model (6) at this scale to

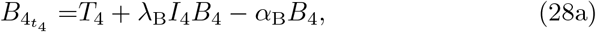

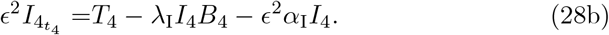

From (28b) we have that *I*_4_ is in a quasi-steady state up to 𝒪(*ϵ*^2^) and therefore,

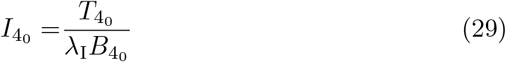

to leading order. As with the *t*_3_ timescale in Section 3.3, the leading order problem will be sufficient to capture the dynamics in this *t*_4_ timescale. Substituting (29) into (28) yields

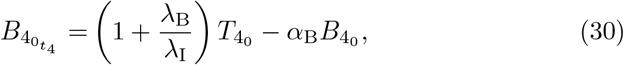

which has solution

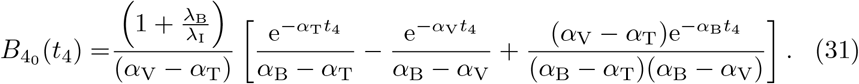

We note that for *t*_4_ ≪ 1 that

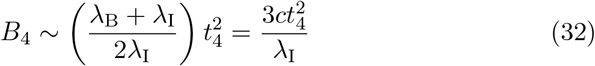

which matches *B*_3_ for *t*_3_ ≫ 1 as required (see (A16) in Appendix A). Following (8), after taking *t* = *ϵ*^−1^*t*_4_ and scaling *A* = *ϵ*^−2^*A*_4_, then the antibody concentration satisfies,

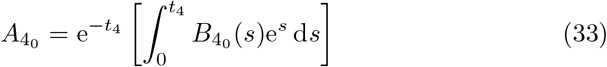

to leading order.

Since we have assumed that *α*_V_, *α*_T_ and *α*_B_ have unique values, for *t*_4_ ≫ 1,

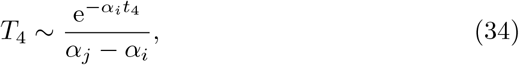

where *α*_*i*_ = min(*α*_V_, *α*_T_) and *α*_*j*_ = max(*α*_V_, *α*_T_). Similarly for plasma B-cells,

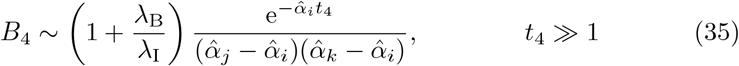

where 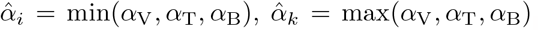, and 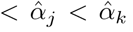. If 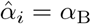 then from (29),

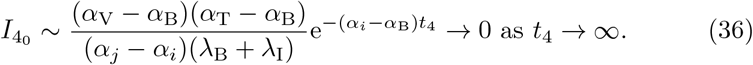

If instead 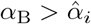 then 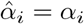 and

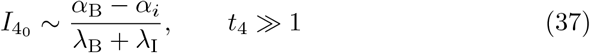

which is a constant. However, if we look at the full equation (28b) then the steady state interleukin concentration, 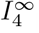, is

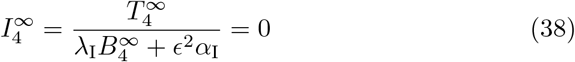

because 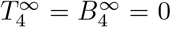. Therefore if *T* and *B* decay at the same rate then a final timescale emerges to resolve the decay of interleukin.

### 3.5 Interleukin Cessation

We assume for this Section that 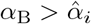 so that (37) is in disagreement with the steady state interleukin (38) when *t*_4_ ≫ 1 necessitating the new timescale. Physiologically, this will occur when plasma B-cells do not decay slower than CD4^+^ T-cells. The failure of interleukin decay stems from neglecting the *α*_I_ term in the denominator of (38) which cannot be done if *B*_4_ ∼ 𝒪(*ϵ*^2^). From (35) this occurs approximately when 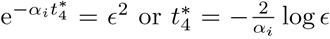 and so we introduce a fifth and final timescale, 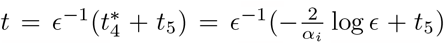 with the additive form of *t*_5_ originating from the exponential decay behaviour in *B*. We substitute this timescale into (7c) for T to yield,

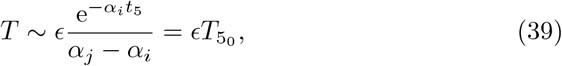

to leading order. The reduced model (6) at this order becomes a fully nonlinear coupled model which cannot be solved analytically. However, from (35) we know that *B*_4_ is decaying exponentially for *t*_4_ ≫ 1. The introduction of the *t*_5_ timescale was to include the natural decay of interleukin and as such we will approximate the plasma B-cell concentration with (35) substituting in the *t*_5_ timescale,

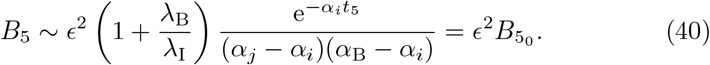

With this approximation, the antibody response can also be continued from (33).

Using (39) and (40) allows simplification of the reduced model (6) to a linear equation for interleukin,

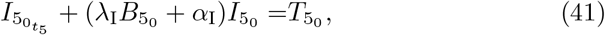

with solution,

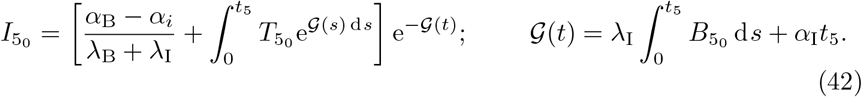

We note that in (42) we have used 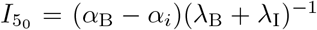 when *t*_5_ = 0 following (37).

The *t*_5_ timescale captures the final decay of interleukin and thus completes the dynamics of the model.

## 4 Results

We begin by demonstrating that the five timescales discussed in Section 3 reasonably capture the model dynamics. We plot *L, V, T, C*, and *F*, which can be determined analytically for all time, in Figure 1. We plot *B, A*, and *I*, which required asymptotic decomposition, in Figure 2. We simulate both the full model (1) and the reduced model (6) to demonstrate the negligible effect of ignoring the parameters *λ*_F_, *λ*_C_, *α*_L_, and *κ*_*i*_. We have chosen *α*_B_ *> α*_V_ *> α*_T_ so that interleukin cessation dynamics can be observed. To indicate the different timescales described in Section 3 we alternate a gray-white background with the first gray background being the *t*_1_ range, the first white background being the *t* ∼ 𝒪(1) range, the next gray background being the *t*_3_ range etc. We plot the time on a logarithmic axis to capture the full range of dynamics from 𝒪(*δ*) to 𝒪(*ϵ*^−1^). In Figure 2, for the asymptotic expansion comparison, we use only the leading order terms that were derived in Section 3. The one exception to this is the *t*_2_ = *t* ∼ 𝒪(1) timescale in Section 3.2 where a two-term expansion was computed to show the loss of asymptotic consistency. Therefore, we use the full two-term expansion for comparison.

**Fig. 1:**
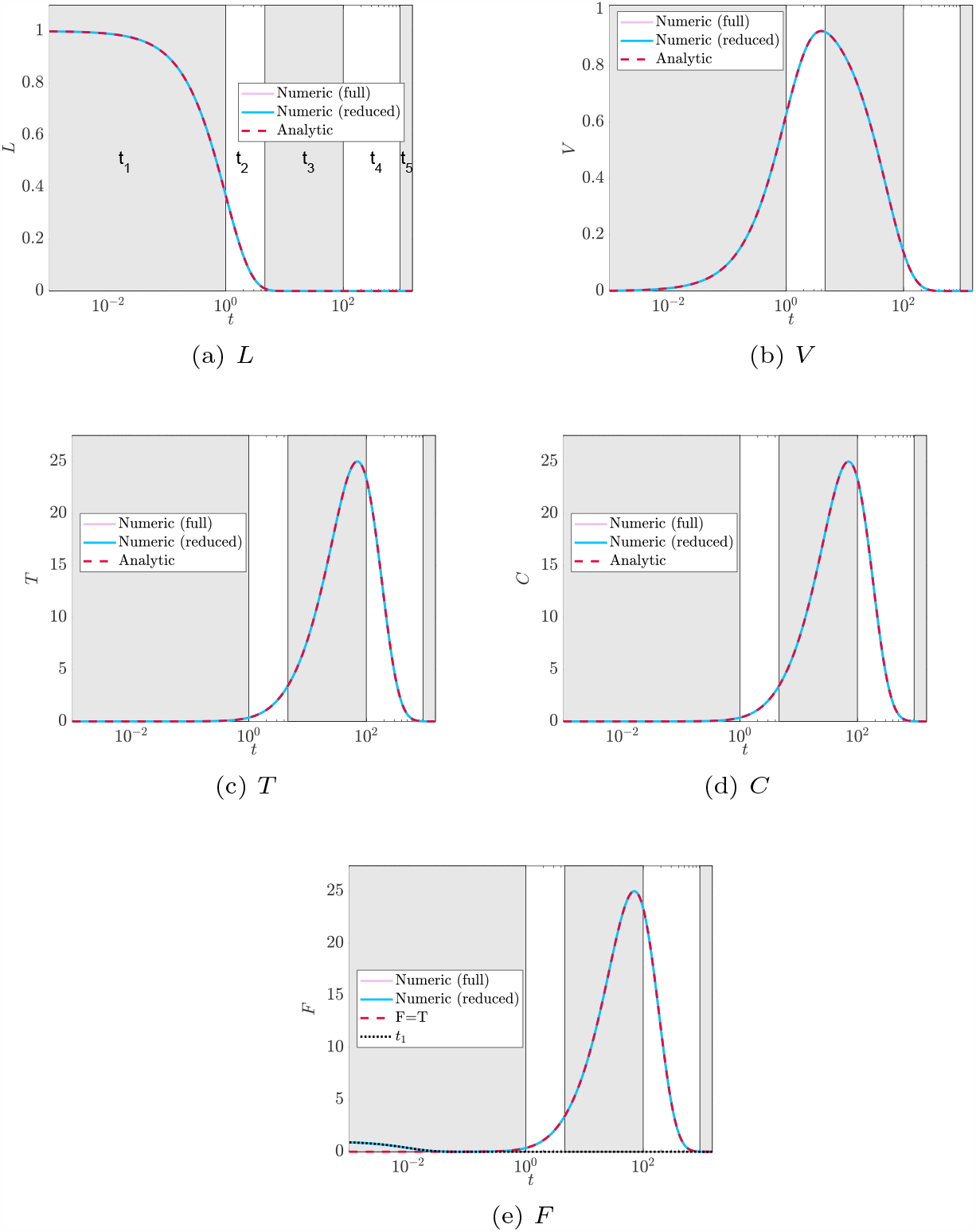
Comparison of the full model (4), the reduced model (6), and the analytic approximations to *L, V, T, C*, and *F* given by (7) and (10) respectively with parameters *ϵ* = *δ* = *κ*_F_ = *κ*_I_ = 0.01, *α*_L_ = *α*_T_ = *α*_C_ = *α*_I_ = *λ*_*i*_ = 1, *α*_V_ = 2, and *α*_B_ = 3 on a logarithmic non-dimensional time series. For all simulations the initial conditions are *L*(0) = *A*(0) = *F*(0) = *I*(0) = 1 and *V* (0) = *T*(0) = *B*(0) = *C*(0) = 0. We alternate the plot with gray and white patches to indicate the regions where each timescale is valid. Since we have taken *ϵ* = 0.01 then the start of each time interval is *t*_1_ = 0.01, *t*_2_ = *t* = 1, *t*_3_ = 4.64, *t*_4_ = 100, and *t*_5_ = 921.03 which are non-dimensional times. For *F* we note the red-dashed curve represents that quasi-steady limit *F* = *T* derived for all timescales beyond *t*_1_ as detailed in section 3.

**Fig. 2:**
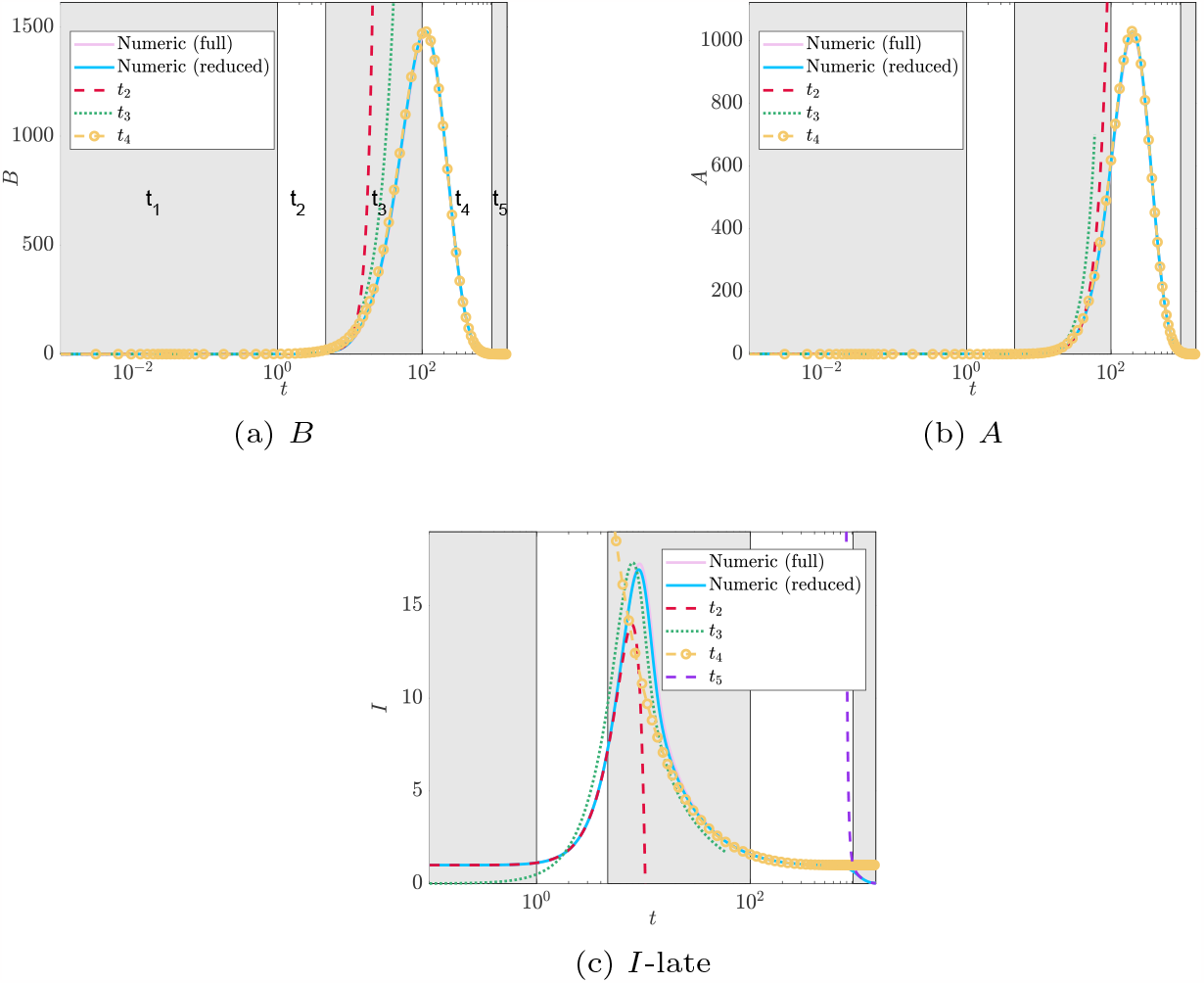
Comparison of the full model (4), the reduced model (6), and the asymptotic approximations to *B, A*, and *I* described in Section 3 with parameters *ϵ* = *δ* = *κ*_F_ = *κ*_I_ = 0.01, *α*_L_ = *α*_T_ = *α*_C_ = *α*_I_ = *λ*_*i*_ = 1, *α*_V_ = 2, and *α*_B_ = 3 on a logarithmic non-dimensional time series. For all simulations the initial conditions are *L*(0) = *A*(0) = *F*(0) = *I*(0) = 1 and *V* (0) = *T*(0) = *B*(0) = *C*(0) = 0. We alternate the plot with gray and white patches to indicate the regions where each timescale is valid. Since we have taken *ϵ* = 0.01 then the start of each time interval is *t*_1_ = 0.01, *t*_2_ = 1, *t*_3_ = 4.64, *t*_4_ = 100, and *t*_5_ = 921.03.

We observe excellent agreement between simulation and analytic results in Figure 1. Since we have chosen a non-zero initial condition for IFN-*γ*, in Figure 1e the initial condition decays to zero in the *t*_1_ timescale before reaching the *F* = *T* quasi-steady value as discussed in Section 3.1. Figure 2 showcases the role of the autocatalytic timescale *t*_3_ discussed in Section 3.3. The plasma B-cell and antibody response in Figures 2a and 2b, respectively, do not have a substantial region in the *t*_3_ timescale where the asymptotic solutions compare favourably with the numerical ones. Instead, the solution transfers smoothly from the *t* ∼ 𝒪(1) timescale (discussed in Section 3.2) to the *t*_4_ timescale (discussed in Section 3.4). However, as seen in Figure 2c, the interleukin dynamics are highly captured by the asymptotic solution in the *t*_3_ timescale and its maximal concentration is reached within the *t*_3_ region. Interleukin plays an activator-inhibitor role in the system. It activates the plasma-B cells to initiate the autocatalytic production but inhibits itself which ultimately terminates the autocatalytic production. The maximal value of interleukin represents a shift from the activator to inhibitor role. As observed in Figure 2c the maximum occurs quite early in the *t*_3_ timescale explaining why *B* and *A* quickly follow the dynamics at the *t*_4_ timescale described in Section 3.4. We emphasize that the solutions in each timescale are formally valid for an arbitrary small parameter, *ϵ*, for the regions they are defined. When choosing an actual value for *ϵ*, such as *ϵ* = 0.01 in Figure 2 then the asymptotic structure can breakdown numerically. This is particularly important for the autocatalytic region described in section 3.3 which occurs at an order *ϵ*^−1*/*3^. Formally, *ϵ*^−1*/*3^ ≫ 1, however, if *ϵ* = 0.01, for example then *ϵ*^−1*/*3^ = 4.6 which is not particularly large. In this timescale we have *I* ∼ 𝒪(*ϵ*^−2*/*3^) ≈ 21 when *ϵ* = 0.01. As can be seen in Figure 2c and is detailed in section 4.1, interleukin transitions through its maximum throughout the *t*_3_ timescale and so very quickly *I* returns to 𝒪(1) practically entering the *t*_4_ regime. Thus, numerical realizations of *ϵ* may cause the solutions in timescales *t*_2_, *t*_3_, and *t*_4_ to be appear to be valid longer (or shorter) than theoretically predicted in the analysis. We could improve the asymptotic agreement by considering the composite solution which is the sum of the different asymptotic solutions with their overlapping contributions removed, but omit the details here.

### 4.1 Maximal Concentrations

The asymptotic solution for interleukin and plasma B-cells at the *t*_3_ timescale provides us a mechanism to determine the maximal interleukin concentration. At the maximal concentration, 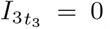, and thus from (23) that 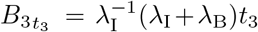 to leading order. Substituting this into the differential equation for *B*_30_ (25) yields,

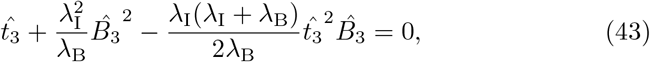

where 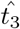 is the time at which the maximal interleukin concentration occurs and we define 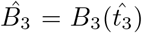. Solving (43) for the plasma B-cell concentration yields,

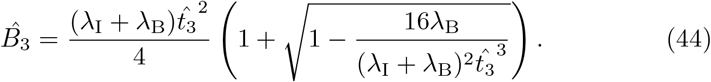

This expression only has a solution so long as 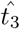 is large enough, namely

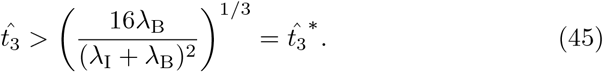

Prior to this time, *B*_3_ cannot grow fast enough to generate an optimal value for the interleukin concentration. Therefore, while (45) does not provide any insight into the value for 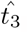, it does provide an estimate for the minimum time before interleukin begins inhibiting antibody production which is useful information for understanding the dynamics of the immune response. To actually estimate the value for 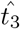, it is more useful to solve (43) for 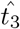. This yields,

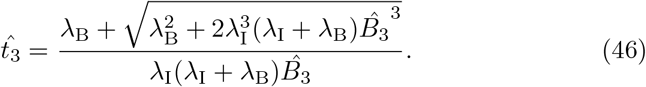

Using (45) as an initial guess, we can compute 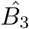 from (26). Substituting this value into (46), we update 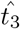. We then iterate until we converge to a desired tolerance. For the parameters in Figure 2c, we have that 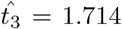 and 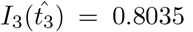. Taking scales into account, these correspond to non-dimensional time 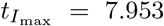 and concentration 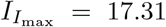 and are comparable to the maximal values numerically obtained from simulating the full model (4), 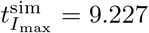 and 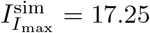 respectively.

The maximum for plasma B-cells occurs in the *t*_4_ timescale. From (30), the maximum *B* concentration to leading order occurs when

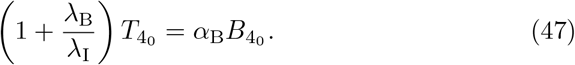

From (27) and (31) this happens when 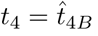, occurring when

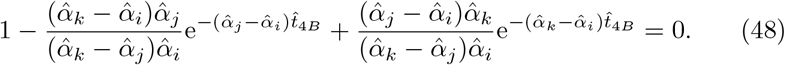

Recall that *α*_V_, *α*_T_, and *α*_B_ are ordered such that 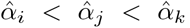. The expression (48) is a polynomial equation in 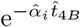 and thus can be solved in a straight forward way. From the parameters in Figure 2a we have that (48) gives 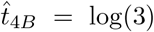 and 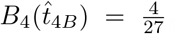. Accounting for scaling, these correspond to non-dimensional time 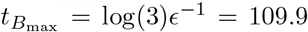 and concentration 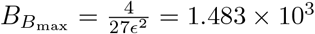 and compare favourably to the numerical values from the simulation of the full model (4) given by 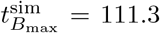 and 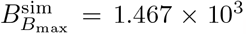. Interestingly, we find that the leading order maximal time given by (48) is independent of the parameters *λ*_B_ and *λ*_I_, depending only on the decay parameters.

The maximum for IgG antibody also occurs in the *t*_4_ timescale and from (6e) occurs at a time 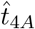 when *A*_4_ = *B*_4_ (with *A*_4_ given by (33) and *B*_4_ given by (31)). Due to the integration in the solution for *A*_4_, this may not be an exponential polynomial (such as is the case with the parameters in Figure 2). However, it can still be solved with a straightforward root-finding process. This yields the time of the maximum antibody concentration for the parameters in Figure 2b as 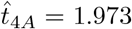 with concentration 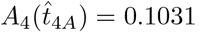.

Accounting for scales, these values are 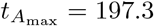 and 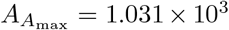 for non-dimensional time and concentration, which compare well to the numerical maximal values from simulating the full model (4), 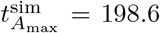 and 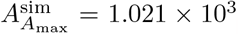. Since the antibody is determined as an integral of the plasma B-cells, its time of maximum concentration also does not depend on the growth parameters *λ*_B_ and *λ*_I_.

### 4.2 Comparison to Data

Parameter fits to the full model (1) for 22 different data sets were performed by Korosec et al (2022) using mixed-effects modelling software Monolix. The data available was for IgG antibodies, interleukin, and IFN − *γ* from patients receiving two doses of SARS-CoV-2 vaccine produced by Pfizer-BioNTech (BNT162b2) or Moderna (mRNA-1273). Each data point refers to a time point measurement of either antibody, IFN-*γ*, or interleukin for a given patient. Since each study has multiple patients, some studies have multiple measurements at a given time. Patients are distinct between studies, but within each study,patients are followed chronologically through time taking additional measurements at different time points. However, these individual patient trajectories cannot be identified from the aggregate data. Therefore, for the purpose of parameter fitting in Korosec et al (2022), “individual” refers to all of the data in a single data set and “population” refers to the collection 22 data sets. For all details about parameter estimates including inferred parameter distributions see Korosec et al (2022) and the supplementary material therein.

Using the fit dimensional parameters from Korosec et al (2022), we compute the equivalent non-dimensional parameters in Table 2 where we also include the average values for each of the two vaccines considered. We bold values in Table 2 that violate the model assumption of parameters being 𝒪(1) or smaller. The scales for each of the vaccine model compartments given by (3) as well as the non-dimensional non-zero initial values 𝒜_0_ and ℐ_0_ are computed in Table 3 for each of the parameter sets (an initial condition ℱ = 0 was taken for all data). We compute the timescales discussed in Section 3 for each of the data sets in Table 4 along with the number of data points that occur within each timescale. The number of data points is important because it provides a measure of data resolution and the ability to capture all the dynamics of the immune response.

**Table 2:**
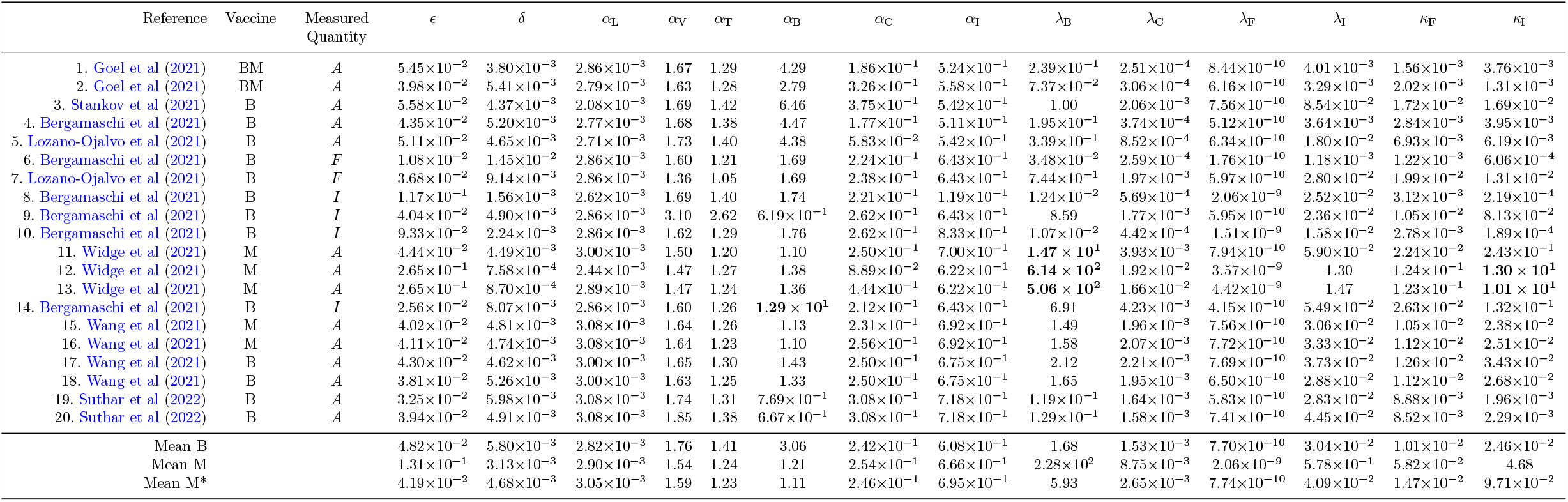
Non-dimensional parameters defined by (5) for various data sets. For the vaccine column B refers to two doses of the vaccine BNT162b2 while M refers to two doses of the vaccine mRNA-1273 (and BM refers to one dose of each). Mean B and Mean M are the average parameter values for BNT162b2 and mRNA-1273 respectively while Mean M* is the average of the mrNA-1273 vaccine with rows 12 and 13 removed. The combined dosage data set from Goel et al (2021) was not considered in the averages. Bolded entries fail the 𝒪(1) assumption of the model (4).

**Table 3:** Scales for each of the variables following (3). All units are arbitrary dependent on the reference with the exception of *F* and *I* which are in units of pg/ml as well as *t* which is in units of days. The values 𝒜 and ℐ are the non-dimensional non-zero initial values for *A* and *I* following (5).

| Reference | Vaccine | Measured Quantity | $V$ | $T$ | $B$ | $A$ | $C$ | $F$ | $I$ | $t$ | $\mathcal{A}$ | $\mathcal{I}$ |
| --- | --- | --- | --- | --- | --- | --- | --- | --- | --- | --- | --- | --- |
| 1. Goel et al (2021) | BM | $A$ | 1 | $9.74 \times 10^{-1}$ | $8.85 \times 10^{-2}$ | $5.69 \times 10^{-1}$ | $2.73 \times 10^{-5}$ | $9.38 \times 10^{-1}$ | 3.76 | 1.30 | 1.02 | $5.70 \times 10^{-1}$ |
| 2. Goel et al (2021) | BM | $A$ | 1 | 1.24 | $8.85 \times 10^{-2}$ | 1.03 | $2.04 \times 10^{-5}$ | 1.21 | 1.31 | $9.26 \times 10^{-1}$ | $5.06 \times 10^{-1}$ | 2.86 |
| 3. Stankov et al (2021) | B | $A$ | 1 | $1.10 \times 10^1$ | 1.17 | 1.71 | $2.79 \times 10^{-5}$ | $1.03 \times 10^1$ | $1.69 \times 10^1$ | 1.16 | $6.55 \times 10^{-1}$ | $1.91 \times 10^{-1}$ |
| 4. Bergamaschi et al (2021) | B | $A$ | 1 | 1.82 | $1.32 \times 10^{-1}$ | 2.55 | $1.85 \times 10^{-5}$ | 1.70 | 3.95 | $9.26 \times 10^{-1}$ | 1.45 | $8.96 \times 10^{-1}$ |
| 5. Lozano-Ojalvo et al (2021) | B | $A$ | 1 | 4.31 | $3.76 \times 10^{-1}$ | 1.72 | $2.34 \times 10^{-5}$ | 4.16 | 6.19 | 1.06 | 1.67 | $5.69 \times 10^{-1}$ |
| 6. Bergamaschi et al (2021) | B | $F$ | 1 | 1.04 | $2.62 \times 10^{-2}$ | $3.18 \times 10^{-1}$ | $5.67 \times 10^{-6}$ | $7.35 \times 10^{-1}$ | $6.06 \times 10^{-1}$ | $2.58 \times 10^{-1}$ | $7.19 \times 10^1$ | 8.81 |
| 7. Lozano-Ojalvo et al (2021) | B | $F$ | 1 | 7.20 | $6.19 \times 10^{-1}$ | 7.37 | $1.93 \times 10^{-5}$ | $1.20 \times 10^1$ | $1.31 \times 10^1$ | $8.77 \times 10^{-1}$ | 3.17 | $3.79 \times 10^{-1}$ |
| 8. Bergamaschi et al (2021) | B | $I$ | 1 | 2.25 | $5.88 \times 10^{-1}$ | 7.41 | $6.67 \times 10^{-5}$ | 1.87 | $2.19 \times 10^{-1}$ | 2.78 | 2.48 | 4.07 |
| 9. Bergamaschi et al (2021) | B | $I$ | 1 | 6.88 | $5.83 \times 10^{-1}$ | 6.24 | $1.92 \times 10^{-5}$ | 6.29 | $8.13 \times 10^1$ | $9.62 \times 10^{-1}$ | 4.21 | $8.00 \times 10^{-3}$ |
| 10. Bergamaschi et al (2021) | B | $I$ | 1 | 1.73 | $3.70 \times 10^{-1}$ | 4.67 | $4.89 \times 10^{-5}$ | 1.67 | $1.89 \times 10^{-1}$ | 2.22 | 5.02 | $1.09 \times 10^1$ |
| 11. Widge et al (2021) | M | $A$ | 1 | $1.38 \times 10^1$ | 1.47 | $2.73 \times 10^1$ | $2.44 \times 10^{-5}$ | $1.35 \times 10^1$ | $2.43 \times 10^2$ | 1.11 | $1.48 \times 10^1$ | $2.04 \times 10^{-2}$ |
| 12. Widge et al (2021) | M | $A$ | 1 | $8.56 \times 10^1$ | $4.18 \times 10^1$ | $8.73 \times 10^2$ | $1.24 \times 10^{-4}$ | $7.42 \times 10^1$ | $1.30 \times 10^4$ | 5.88 | $5.04 \times 10^{-1}$ | $3.99 \times 10^{-4}$ |
| 13. Widge et al (2021) | M | $A$ | 1 | $7.49 \times 10^1$ | $4.40 \times 10^1$ | $8.52 \times 10^2$ | $1.53 \times 10^{-4}$ | $7.38 \times 10^1$ | $1.01 \times 10^4$ | 5.88 | $6.06 \times 10^{-1}$ | $3.40 \times 10^{-4}$ |
| 14. Bergamaschi et al (2021) | B | $I$ | 1 | $1.64 \times 10^1$ | 1.10 | $1.41 \times 10^1$ | $1.34 \times 10^{-5}$ | $1.58 \times 10^1$ | $1.32 \times 10^2$ | $6.10 \times 10^{-1}$ | 1.56 | 1.79 |
| 15. Wang et al (2021) | M | $A$ | 1 | 6.56 | $6.62 \times 10^{-1}$ | 9.51 | $2.27 \times 10^{-5}$ | 6.33 | $2.38 \times 10^1$ | 1.03 | 5.35 | $2.19 \times 10^{-1}$ |
| 16. Wang et al (2021) | M | $A$ | 1 | 6.92 | $7.21 \times 10^{-1}$ | $1.05 \times 10^1$ | $2.32 \times 10^{-5}$ | 6.73 | $2.51 \times 10^1$ | 1.05 | 4.82 | $2.04 \times 10^{-1}$ |
| 17. Wang et al (2021) | B | $A$ | 1 | 7.80 | $8.30 \times 10^{-1}$ | $1.22 \times 10^1$ | $2.37 \times 10^{-5}$ | 7.55 | $3.43 \times 10^1$ | 1.08 | 4.16 | $1.51 \times 10^{-1}$ |
| 18. Wang et al (2021) | B | $A$ | 1 | 6.86 | $6.40 \times 10^{-1}$ | 9.60 | $2.00 \times 10^{-5}$ | 6.71 | $2.68 \times 10^1$ | $9.52 \times 10^{-1}$ | 5.32 | $1.99 \times 10^{-1}$ |
| 19. Suthar et al (2022) | B | $A$ | 1 | 5.48 | $5.02 \times 10^{-1}$ | 4.89 | $1.75 \times 10^{-5}$ | 5.33 | 1.96 | $8.33 \times 10^{-1}$ | $2.13 \times 10^1$ | 3.17 |
| 20. Suthar et al (2022) | B | $A$ | 1 | 5.28 | $6.94 \times 10^{-1}$ | 7.47 | $2.22 \times 10^{-5}$ | 5.11 | 2.29 | 1.01 | $1.58 \times 10^1$ | 3.19 |

**Table 4:**
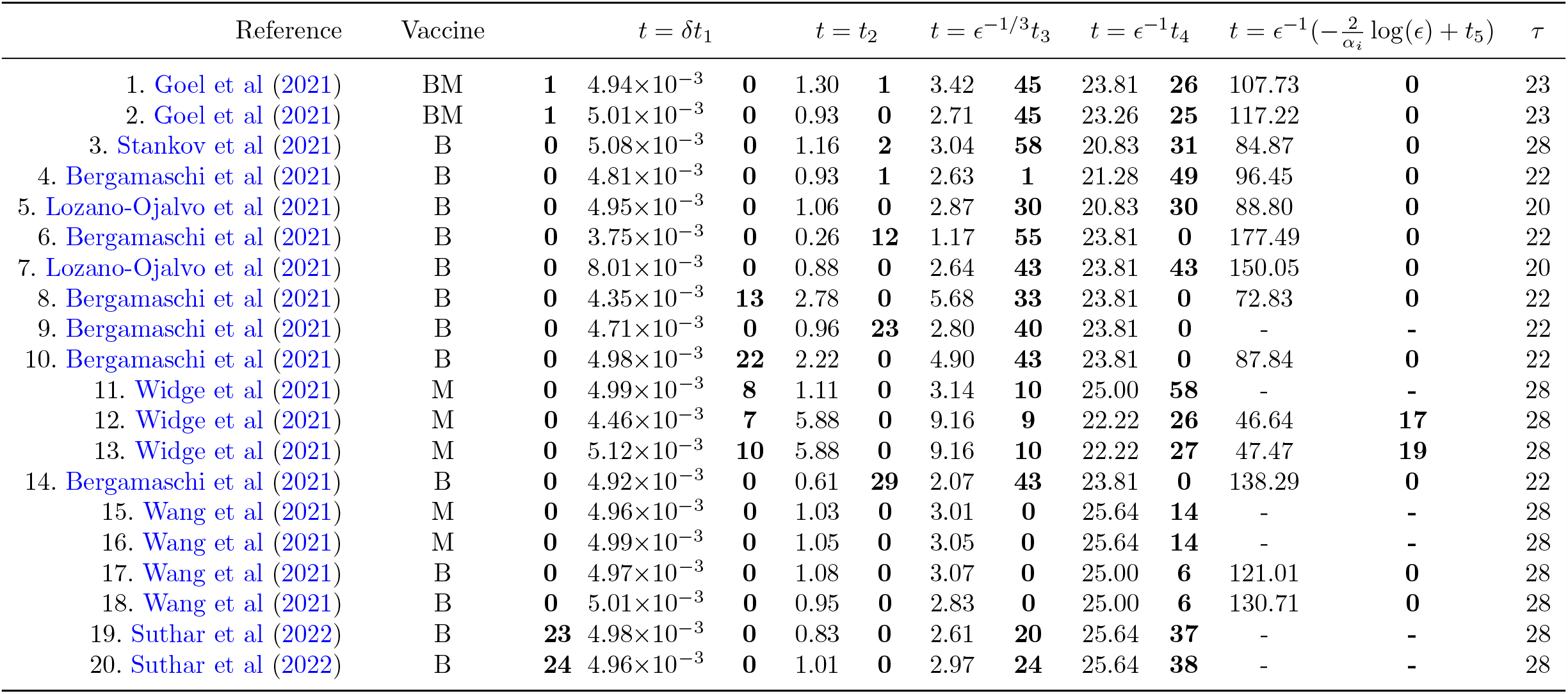
Values for the start of each of the timescales (in days) discussed in Section 3 with *ϵ* taken from Table 2 and *t* determined as 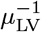 calculated in Korosec et al (2022). The bold values are the number of data points in each data set that lie between timescales, i.e. *t*_*i*_ *< t < t*_*i*+1_ and the first column of bold values are those point in 0 ≤ *t < t*_1_. A dash in the *t*_5_ column indicates that min(*α*_V_, *α*_T_, *α*_B_) = *α*_B_ and thus that interleukin cessation does not occur. The final column *τ* is the time (in days) that the second dose of vaccine was administered in each study.

The number of data points in Table 4 is slightly misleading. Many data sets have multiple patients and therefore much of the data is taken at a single time point for multiple people rather than as a time series for a single person. Secondly, the number of data points in 0 ≤ *t < t*_1_ (first column of bold values) are all initial values taken just after vaccination (day 0) and do not actually capture any of the fast IFN-*γ* dynamics discussed in Section 3.2. The model analysis predicts that the majority of antibody activity occurs within the *t*_3_ and *t*_4_ timescales and this is precisely where the majority of data points are collected. Furthermore, we remark the onset of the *t*_4_ timescale which is the timescale over which antibody concentrations peak and decay aligns remarkably well with the timing of the second dosage given as the last column of Table 4.

Table 5 calculates an estimation of the maximal concentration and times they occur for interleukin, plasma B-cells, and IgG antibody following the discussion in Section 4.1. The sequence of timescales discussed in Section 3 indicates that interleukin reaches maximal value first (in the *t*_3_ timescale) which triggers the inhibition process of plasma B-cells leading to its maximal value (in the *t*_4_ timescale) and then the maximal value of antibody followed shortly. Thus, generally we suspect an ordering of 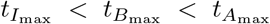 which holds in Table 5 except for data sets 8 and 10 from Bergamaschi et al (2021). These values have been indicated in bold in Table 5.

**Table 5:** Maximal concentrations of interleukin, plasma B-cells, and IgG antibody and the times that they occur (in days). The time 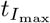 is determined from solving (46), the time 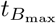 from solving (48), and 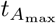 is the time when *A*_4_ = *B*_4_. The maximal concentrations are given by substituting the optimal times into the respective asymptotically determined concentrations given in Section 3. The time 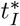 is given by (45). The times should be sequential in that 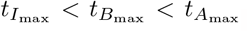. Bold values in the table represent data sets where this is violated.

| Reference | Vaccine | $t_I^*$ | $t_{I_{\max}}$ | $I_{I_{\max}}$ | $t_{B_{\max}}$ | $B_{B_{\max}}$ | $t_{A_{\max}}$ | $A_{A_{\max}}$ |
| --- | --- | --- | --- | --- | --- | --- | --- | --- |
| 1. Goel et al (2021) | BM | 4.02 | 15.65 | 31.28 | 23.07 | $9.81 \times 10^1$ | 43.17 | $4.21 \times 10^2$ |
| 2. Goel et al (2021) | BM | 5.84 | 16.96 | 19.17 | 25.94 | $1.04 \times 10^2$ | 45.80 | $8.38 \times 10^2$ |
| 3. Stankov et al (2021) | B | 2.38 | 7.50 | 36.51 | 17.41 | $1.70 \times 10^2$ | 34.68 | $1.58 \times 10^2$ |
| 4. Bergamaschi et al (2021) | B | 4.29 | 12.77 | 36.84 | 19.80 | $1.91 \times 10^2$ | 37.45 | $2.42 \times 10^3$ |
| 5. Lozano-Ojalvo et al (2021) | B | 3.49 | 10.62 | 31.32 | 19.25 | $1.43 \times 10^2$ | 36.39 | $4.28 \times 10^2$ |
| 6. Bergamaschi et al (2021) | B | 7.55 | 9.58 | 15.64 | 32.06 | $8.24 \times 10^2$ | 53.33 | $7.38 \times 10^3$ |
| 7. Lozano-Ojalvo et al (2021) | B | 2.71 | 7.74 | 42.73 | 35.55 | $1.86 \times 10^3$ | 57.66 | $1.69 \times 10^4$ |
| 8. Bergamaschi et al (2021) | B | 5.20 | **35.76** | 2.23 | **29.68** | 6.75 | **50.31** | $6.11 \times 10^1$ |
| 9. Bergamaschi et al (2021) | B | 1.23 | 4.35 | 84.03 | 27.18 | $9.34 \times 10^3$ | 50.23 | $7.24 \times 10^4$ |
| 10. Bergamaschi et al (2021) | B | 6.24 | **34.92** | 2.53 | **30.87** | 8.03 | **51.84** | $7.38 \times 10^1$ |
| 11. Widge et al (2021) | M | 1.03 | 3.98 | 165.69 | 39.82 | $3.19 \times 10^4$ | 63.39 | $4.59 \times 10^5$ |
| 12. Widge et al (2021) | M | 0.30 | 3.48 | 806.85 | 32.54 | $4.08 \times 10^4$ | 52.80 | $6.45 \times 10^5$ |
| 13. Widge et al (2021) | M | 0.32 | 3.64 | 685.67 | 32.91 | $3.21 \times 10^4$ | 53.25 | $4.71 \times 10^5$ |
| 14. Bergamaschi et al (2021) | B | 1.32 | 3.24 | 133.81 | 18.82 | $4.23 \times 10^3$ | 39.66 | $3.53 \times 10^4$ |
| 15. Wang et al (2021) | M | 2.17 | 7.36 | 56.01 | 38.71 | $3.11 \times 10^3$ | 62.53 | $3.42 \times 10^4$ |
| 16. Wang et al (2021) | M | 2.13 | 7.31 | 56.66 | 39.28 | $3.24 \times 10^3$ | 63.23 | $3.65 \times 10^4$ |
| 17. Wang et al (2021) | B | 1.94 | 6.75 | 65.97 | 34.46 | $3.31 \times 10^3$ | 56.89 | $3.63 \times 10^4$ |
| 18. Wang et al (2021) | B | 2.11 | 6.77 | 60.90 | 35.96 | $3.58 \times 10^3$ | 58.73 | $4.05 \times 10^4$ |
| 19. Suthar et al (2022) | B | 4.45 | 11.61 | 12.41 | 42.47 | $4.34 \times 10^2$ | 67.51 | $3.35 \times 10^3$ |
| 20. Suthar et al (2022) | B | 4.09 | 12.17 | 11.86 | 42.79 | $3.03 \times 10^2$ | 68.29 | $2.60 \times 10^3$ |

We plot the data of each reference data set, along with the full simulated model (4) and each of the asymptotic approximations, in Figure 3 for antibody data and Figure 4 for IFN-*γ* and interleukin data. As with Figure 1 and 2, we alternate gray-white backgrounds to showcase the different timescales. We plot the asymptotic solutions in each of the regions they are valid, but only include the time regions up to the terminal data point in each study. Based on the discussion surrounding Figure 2c about practical asymptotic consistency, we extend the asymptotic solutions for *t*_2_ and *t*_4_ into the *t*_3_ regime because the practical validity can be different than what is theoretically predicted. In Figure 4c and 4e, we do not include the *t*_4_ asymptotic solution in the *t*_3_ region because it is outside the plotting window. In Figures 3 and 4 we also plot three vertical dashed lines which correspond to the maximal times (in chronological order) for each of the concentrations *I, B*, and *A* given in Table 5. If less than three lines are present it means that the predicted optima occur outside the time window where data was available.

**Fig. 3:**
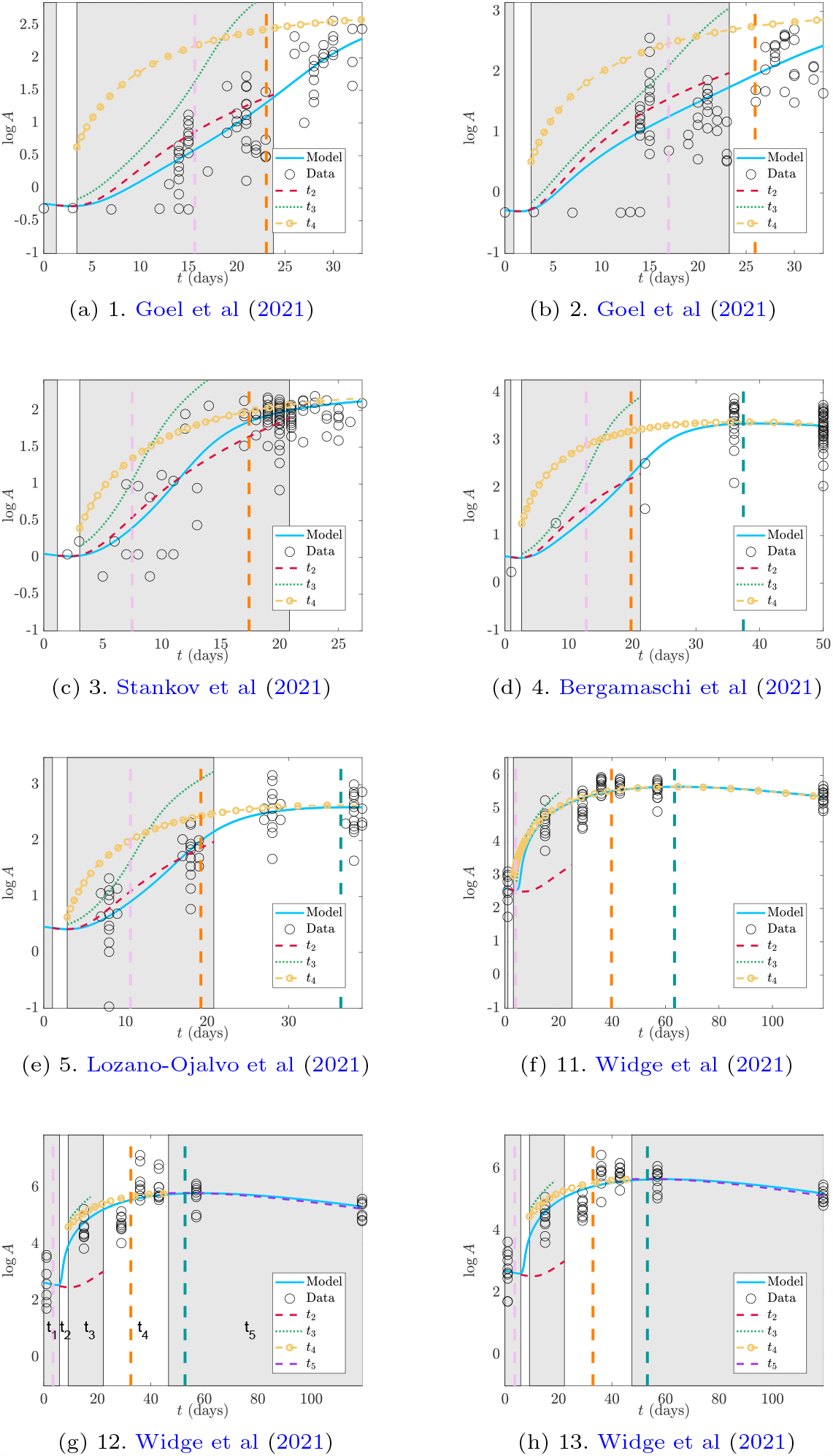
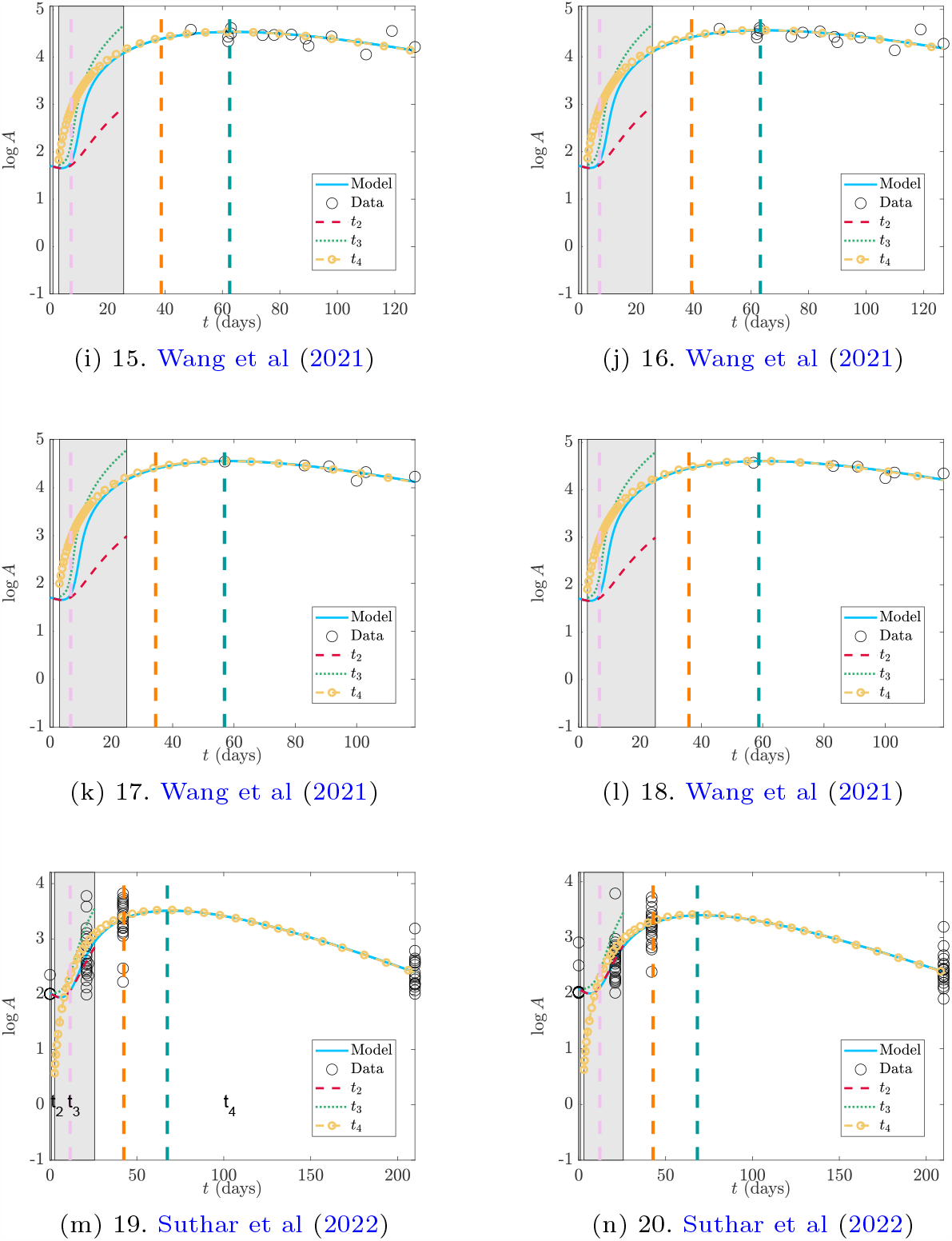
Comparison of the full model (4), the analytic approximations described in Section 3 and data from the cited reference for IgG antibody *A*. Parameters and initial values are listed in Table 2 and Table 3 respectively (with corresponding number entry). The alternating patches are the timescale windows in Table 4. The vertical dashed lines are the times for the maximal concentrations in interleukin (pink), plasma B-cells (orange), and antibody (teal) from Table 5 ordered chronologically. We note the scales on the axes are different for some sub figures.

**Fig. 4:**
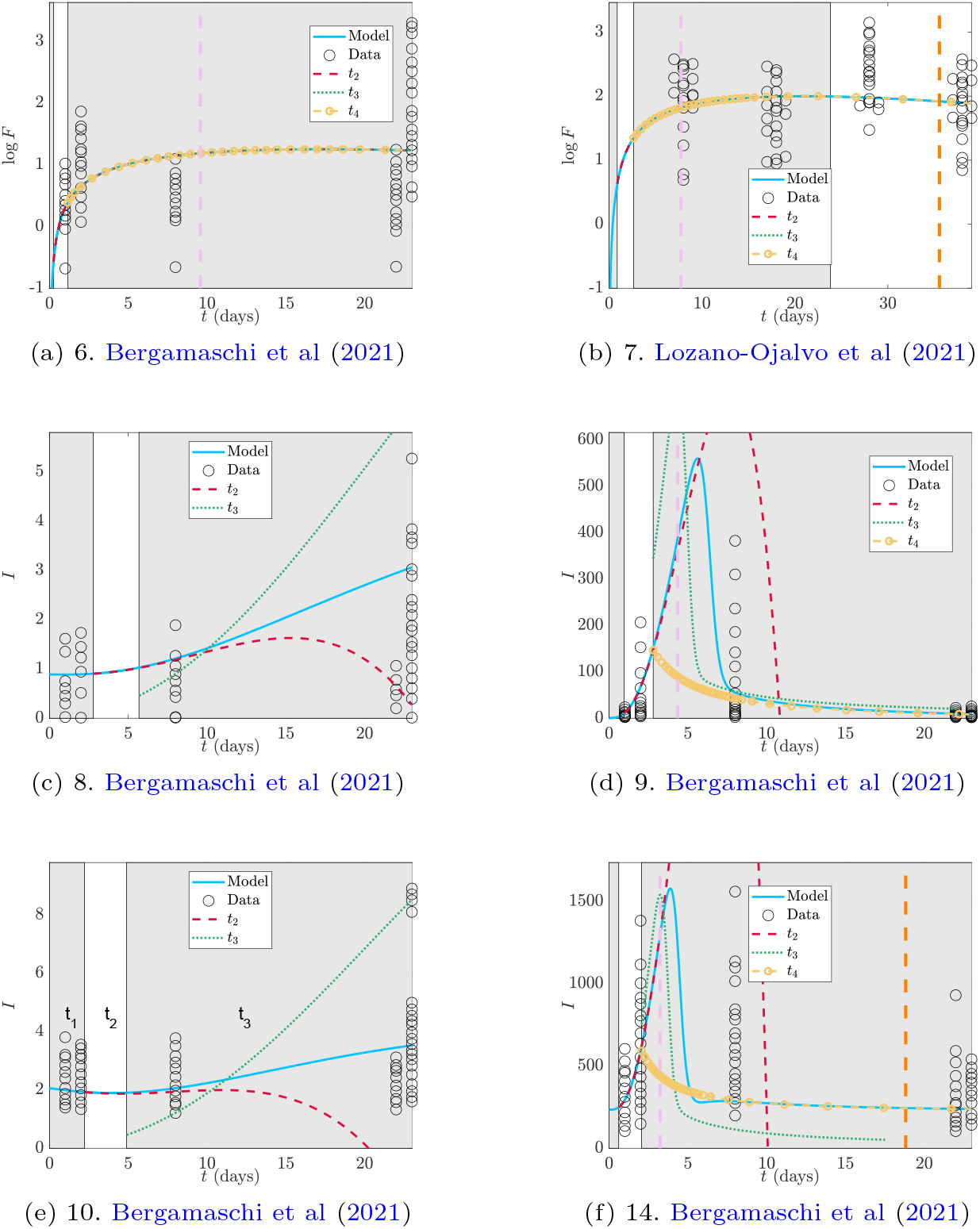
Comparison of the full model (4), the analytic approximations described in Section 3 and data from the cited reference for IFN-*γ* (*F*) and interleukin (*I*). Parameters and initial values are listed in Table 2 and Table 3 respectively (with corresponding number entry). The alternating patches are the timescale windows in Table 4. The vertical dashed lines are the times for the maximal concentrations in interleukin, plasma B-cells, and antibody from Table 5 ordered chronologically. We note the scales on the axes are different for some sub figures.

### 4.3 Multiple Doses

We note that the original model (1) as posed does not allow for multiple doses which is seemingly problematic as all patient data used in Section 4.2 does include a second dose with second dosage times given in Table 3. Mathematically, it is straight-forward to account for additional doses in the model by including impulse terms to the LNP compartment of the model. However, when we compare the model with one and two doses to the data we seem to get a paradoxical conclusion that the one-dose model fits better despite the patient having received two doses (see Figure 5 for an example). One possible explanation is that the parameters fit in Korosec et al (2022) have accounted for a second dose through fitting. Furthermore, the second dosage time is in the *t*_4_ timescale where the peak of antibody occurs, thus creating a potential resolution issue. However, since the second dose does not affect the immune response timescales, nor the general analysis completed here, we save any further discussion of multiple doses for future work.

**Fig. 5:**
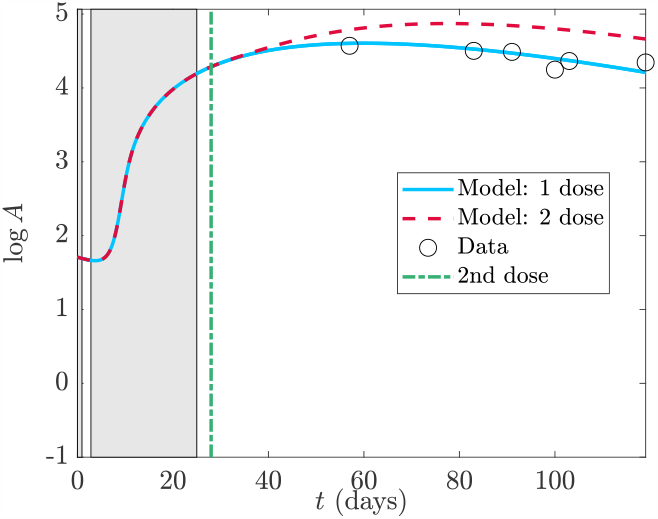
Comparison of data from Wang et al (2021) to the full model (4) with 1 or 2 doses using the parameters of data set 18 in Tables 2, 3 and second dosage time in Table 4. The alternating patches are the timescale windows in Table 4

## 5 Discussion

During the model formulation in Section 2.1 we identified parameters *λ*_F_, *λ*_C_, and *α*_L_ as likely being unidentifiable due to very small pre-factors and hence neglected them in the reduced model (6). Table 2 confirms this hypothesis as each of these parameters are very small based on fit data. We further set each of the saturation parameters to zero under the notion that we would not be over-saturated in plasma B-cells and interleukin. These parameters are indeed generally small in Table 2 justifying the assumption.

A further assumption in the non-dimensional model (4) was that the parameters are 𝒪(1) or smaller so that the model is sensibly scaled. We note that this is mostly true in Table 2 with exceptions to this noted in bold. Data set 14 from Bergamaschi et al (2021) has an anomalous value for *α*_B_. From Figure 4f we observe that most of the data is concentrated around the peak with some later data corresponding to the second dose time. Since *α*_B_ is a decay parameter it will have the most significant contribution on interleukin after the peak value. Furthermore, from Section 3.4 the quasi-steady interleukin concentration (37) depends linearly on *α*_B_, however there is very little data in this region to inform the parameter. Therefore, the value of *α*_B_ in data set 14 may be a consequence of poor data fidelity.

The more interesting anomalous parameters are those for *λ*_B_ (data sets 11, 12, and 13) and *κ*_I_ (data sets 12 and 13) as these all belong to data collected by Widge et al (2021) for the mRNA-1273 vaccine. The data in this paper measured antibody response for various ages. Data set 11 is for ages 18-55, data set 12 ages 56-70, and data set 13 ages 70 and over. We see a dramatic difference between those under and over the age of 55. While age effects on response were discussed by Korosec et al (2022), the parameter contrast was not as apparent in the dimensional form. *λ*_B_ and *κ*_I_ are associated with the autocatalytic production of plasma-B cells and ultimately antibodies. A higher value leads to more rapid production which due to the inhibiting effects on interleukin can also lead to quicker decay.

We note in Table 4 that the *t*_3_ timescale representing the onset of autocatalytic production starts several days later in the older-patient data sets of Widge et al (2021) than the younger-patient data set, but the *t*_4_ timescale representing peak antibody starts a few days sooner. Therefore, overall, the time window over which antibody levels are high is much narrower for ages 56 and greater compared to those below this age. We also note that the youngest age group has *α*_B_ as the slowest decay parameter compared to the two older data sets and thus does not have predicted interleukin cessation. There is no confirmation for this in the data set because interleukin data was not recorded for these cohorts.

Interestingly, the predicted maximal antibody concentrations from Table 5 are higher for the two older-age data sets of Widge et al (2021) compared to the data set for those under 56 with the highest maximum concentration predicted in the 56-70 data set. However, the decay rates *α*_B_ and *α*_T_ also increase, as observed in Table 2 and therefore a high concentration will also experience rapid decay, potentially explaining this counter intuitive result and providing insight into the body’s immunosenescence. We find that *λ*_B_ and *λ*_I_ seem to be age-dependent. These parameters dictate how well the body transitions from vaccine priming (the adapted immunity response) to self-production of antibody precursors (the innate immune response) and therefore provide detail on the immunogenicity of the vaccine. Overall, the analysis indicates the existence of, and possible mechanism for, an age dependent vaccine and natural immune response. A reduced vaccine efficacy with age has been reported in studies such as by Menni et al (2022).

The anomalous bold values in Table 2 also skew the mean values for the mRNA-1273 vaccine making it appear as if there is discernible difference between that and the BNT162b2 vaccine. However, if data sets 12 and 13 of Widge et al (2021) are removed then the new mean for vaccine mRNA-1273 (Mean M* in Table 2) is much more comparable with the mean for BNT162b2, indicating that there is no appreciable difference between the vaccines in terms of immune response parameters. It may appear in Table 5 that there are discernible differences in peak antibody levels between the two vaccines as computing the means for the two vaccines shows a nearly 20 fold difference between the two. However, this comparison cannot be made because the antibody concentrations that are reported are in arbitrary units and may be different for each study. While it is omitted here, redoing the analysis on the maximal antibody concentration in dimensionless units shows approximately a two-fold difference which makes them more comparable.

The data sets of Suthar et al (2022) separated antibody response by sex with data set 19 being male and data set 20 being female. We see, both from the parameters in Table 2 and the timescales in 4, that there is no discernible difference between sexes in the vaccine response.

Interestingly, aside from the two over-55 groups of Widge et al (2021), none of the mRNA-1273 studies considered here have interleukin cessation, whereas all of the BNT162b2 studies do with the exception of those by Suthar et al (2022). However, it is worth noting that the studies by Suthar et al (2022) also have data at the latest time points of all of the studies considered. Overall, the late-time study of interleukin concentration in each of the mRNA-1273 and BNT162b2 vaccines would be an interesting exploration.

Non-anomalous *λ*_B_ values in Table 2 are almost all 𝒪(1). The three smallest values are of order 𝒪(10^−2^) and belong to IFN-*γ* data set 6 and interleukin data sets 8 and 10. It is not surprising that IFN-*γ* parameter fitting may not capture a representative value of *λ*_B_ as the profile entirely mimics that of the CD4^+^ T-cell population whose dynamics do not depend on *λ*_B_. As for the interleukin data sets, observing Figures 4c and 4e, we note that the graphs indeed look strange. Unlike the other interleukin data, there is no early peak, likely due to influence from the later data when the second dose is administered. Figures 4d and 4f where *λ*_B_ ∼ 𝒪(1) do have early peaks in the model simulation. While we do not plot it here, changing the value of *λ*_B_ for data sets 8 and 10 to be of 𝒪(1) indeed introduces a peak into the model consistent with the other data sets. Another indication that the fitting to data sets 8 and 10 is problematic is observed in Table 5 where for those particular data sets we have the anomalous result that 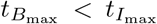 counter to the other data sets and overall model analysis. Overall, the scaling analysis allows us to interrogate parameters far from the mean and analyze their validity.

Unlike the parameter *λ*_B_, the values of *λ*_I_ are much smaller than 𝒪(1) generally, except for the anomalous data sets 12 and 13 previously discussed. Even though the analysis assumed *λ*_I_ ∼ 𝒪(1) the fact that *λ*_I_ ≪ 1 does not impact the overall immune response, it just creates some additional complexity between timescales *t*_3_ and *t*_4_. Specifically, if *λ*_B_ ∼ 𝒪(1) and *λ*_I_ ≪ 1 then when the *t*_3_ timescale begins, the plasma B-cell concentration is very high but the interleukin concentration remains small because of the weak non-linear decay. As such, the plasma B-cell concentrations grow explosively during which a new timescale 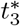 will emerge. This timescale separates from *t*_3_ similar to the interleukin cessation scale, and the concentration of plasma B-cells will become large enough that the weak interleukin inhibition becomes dominant. Overall, the interleukin activator-inhibitor dynamics proceed sequentially from activation to inhibition rather than concurrently as was presented here. Since the overall behaviour is captured by the limit we have chosen here, we omit details of the more general case.

Many of the insights discussed on data fidelity and parameter estimates are due to the explicit analytic expressions obtained. These expressions also provide biological insight. For example, through (45) we understand the minimum time for interleukin to begin inhibiting antibody production and that it depends on two dimensionless groupings, *λ*_B_ and *λ*_I_. We discussed that the peak antibody response seems to highly correlate with age and the result of (45) provides a reduced set of parameters to investigate age dependence in. Our study also showed the delicate relationship that each of the decay parameters have to long-term immune behaviour. If B-cells decay quickest so that *α*_B_ is the largest decay parameter then quasi-steady interleukin concentrations can persist for a long time. Otherwise, interleukin decays quickly. This means that interleukin concentration data at long time points after vaccination can provide insight into the whether the system is dominated by T-cell or B-cell decay.

### 5.1 Conclusions and Future Work

Overall, we have presented a model for LNP delivered mRNA vaccines originally derived in Korosec et al (2022). Through non-dimensionalisation we were able to reduce the model and identify five distinct timescales where different parts of the immune response dominated. Comparing our model to data we were able to identify data sets with different immune response, namely those associated with age, and also identify parameter sets which significantly differ from the mean.

Identifying important timescales of the immune response dynamics provides a framework for clinical data collection. The timescales are determined from two parameters, the rate of vaccine conversion in cells, *μ*_LV_, and the decay rate of IgG antibodies, *γ*_A_. If those can be measured independently then timescales can be estimated and clinicians have windows with which to collect data. This avoids unnecessary sampling and missing important dynamics. Furthermore, it increases the likelihood of capturing data related to peak concentrations which can lead to stronger parameter estimation and thus stronger model prediction. If estimates of *μ*_LV_ and *γ*_A_ are not available because of novel or emerging diseases, we recommend a two-stage trial. The first will create coarse window estimates based on parameters from comparative diseases. Parameter estimation will be done on these windows to refine estimates of the timescales at which point the refined trials discussed above can be conducted.

## Data Availability

All data produced are available online at https://github.com/ionmoles/mRNA-inhost

## Acknowledgments

The authors are grateful to James Watmough, Lindi Wahl, David Dick, and Samaneh Gholami for fruitful discussions during the completion of this manuscript.

## Declarations

### Funding

This work was supported by Natural Sciences and Engineering Research Council of Canada (NSERC) Discovery grants (IRM, JMH) an NSERC Emerging and Infectious Disease Modelling Initiative (IRM, JMH), and the NSERC postdoctoral fellowship program (CSK).

### Competing Interests

The authors declare no competing interests

### Ethics Approval

Not applicable

### Consent to participate

Not applicable

### Consent for publication

Not applicable

### Availability of data and materials

The datasets generated during and/or analysed during the current study are available in the GITHUB repository, https://github.com/ionmoles/mRNA-inhost

### Code availability

The code generated during the current study are available in the GITHUB repository, https://github.com/ionmoles/mRNA-inhost

## Authors’ contributions

All authors devised the study and performed the literature review. IRM did the primary analysis and coding. All authors edited the manuscript and provided feedback.

## Appendix A Plasma B-cell concentration in the *t*_3_ Timescale

The autocatatlyic problem is given by (20) in Section 3.3 with timescale *t* = *ϵ*^−1*/*3^*t*_3_. We pose an expansion *B*_3_ ∼ *B*_30_ + *ϵ*^1*/*3^*B*_31_ where the leading order problem is given by (25) in Section 3.3,

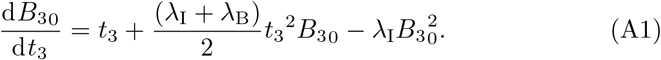

As this is a non-linear Riccati equation we can simplify using the transformation 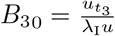 which allows us to write (A1) as

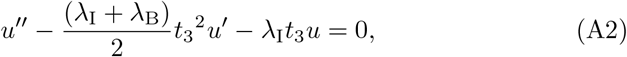

where prime indicates differentiation with respect to *t*_3_. Further transforming *u* = *t_3_w*(*z*) with 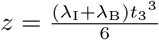 yields,

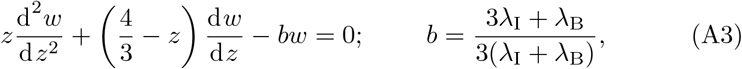

which is Kummer’s differential equation and has as solutions Kummer’s functions M(*x, y, z*) and U(*x, y, z*) (see Kummer (1837); Tricomi (1947); Slater (1960); Polyanin and Zaitsev (2017)),

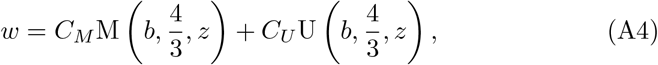

for arbitrary constants *C*_*M*_ and *C*_*U*_. Therefore, *B*_30_ has solution,

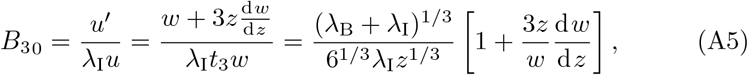

which from (15a) must satisfy 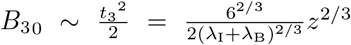 for *z* ≪ 1. Substituting (A4) into (A5) and expanding for *z* ≪ 1 yields

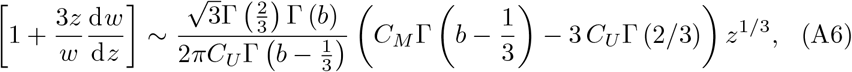

to leading order where Γ(*z*) is the usual Gamma function (see for example Polyanin and Zaitsev (2017)). (A6) does not match the required form of *B*_3_ for *z* ≪ 1 and therefore we must choose *C*_*U*_ so that this term is zero,

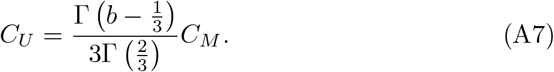

Returning to the expansion (A6) the next order term is

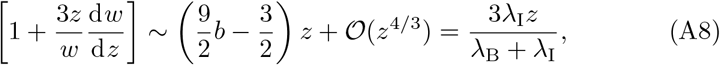

where we have used the definition of *b* from (A3). Substituting this into (A5) we get that to leading order *B*_30_ satisfies,

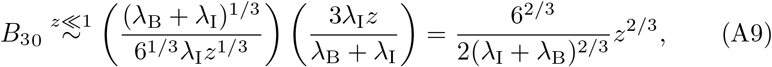

which satisfies the correct matching condition for all *C*_*M*_. Therefore, without loss of generality we can take *C*_*M*_ =1. Overall then, the solution for *B*_30_ satisfies

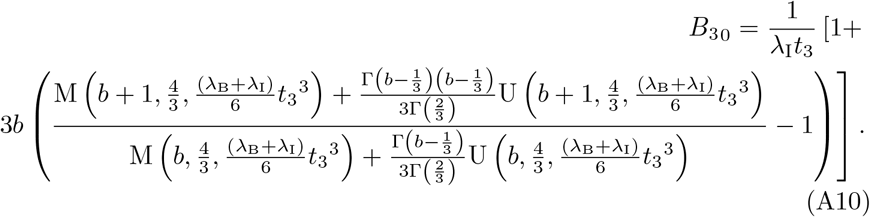

### A.1 No Loss of Asymptotic Consistency

The next order problem of (20) is

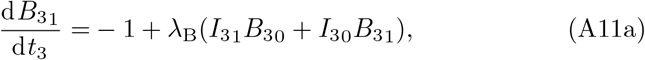

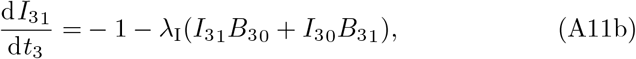

where once again multiplying (A11a) by *λ*_I_ and (A11b) by *λ*_B_ and adding produces a reduced equation

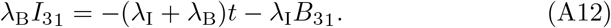

Substituting this into (A11a) and recognizing from (22) that 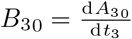 yields an equation for *B*_31_,

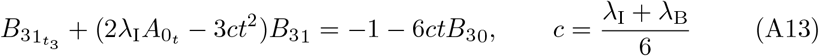

which is linear and has solution,

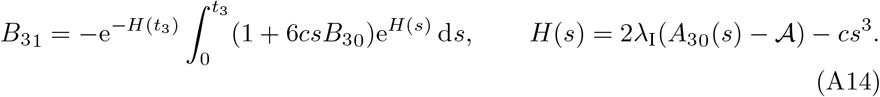

We already know from the solution for the CD4^+^ T-cells in the *t*_3_ timescale given by (19) that an important timescale emerges when *t*_3_ ∼ 𝒪(*ϵ*^−2*/*3^) (*t* ∼ *ϵ*^−1^). However, in Section 3.2 the solution for *B, A*, and *I* broke down sooner thus introducing the *t*_3_ timescale. It is therefore important to investigate any additional timescales that may emerge between *t* ∼ 𝒪(1) and *t* ∼ 𝒪(*ϵ*^−1^). To do so we investigate *B*_30_ and *B*_31_ for *t*_3_ ≫ 1.

The asymptotic expansions of the Kummer functions for large argument are (see for example Abramowitz and Stegun (1983); Andrews et al (1999); Polyanin and Zaitsev (2017)),

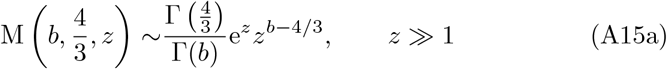

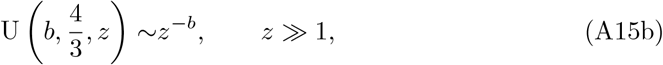

and so from (26) for *t*_3_ ≫ 1 we have that

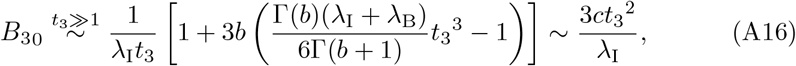

and also from (22) that,

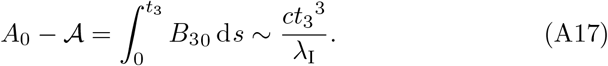

With these asymptotic expansions then, from (A14), we have that,

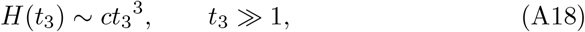

and also that

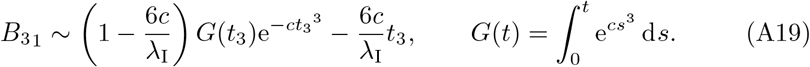

*G*(*t*) can be written in terms of an incomplete Gamma function (see Polyanin and Zaitsev (2017)) and thus the first term in (A19) vanishes for *t*_3_ ≫ 1. Thus, overall we have that

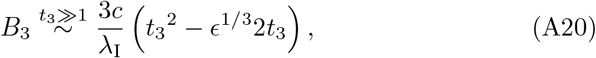

and *B*_3_ does not lose asymptotic consistency at least up to 𝒪(*ϵ*^1*/*3^) since *t*_3_ *< t*_3_^2^ for *t*_3_ ≫ 1. Similarly from (21b) and (A11b) we have the expressions for interleukin under *t*_3_ ≫ 1 satisfy *I*_30_ = *I*_31_ = 0 and therefore

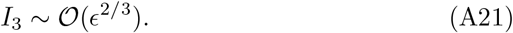

Thus, interleukin also does not lose asymptotic consistency up to the order considered. Finally from (8), since *A* is bounded from above by the integral of *B* then since *B* does not lose asymptotic consistency then neither does *A*.

Therefore, there are no additional intermediate timescales between *t* ∼ 𝒪(*ϵ*^−1*/*3^) and *t* ∼ 𝒪(*ϵ*^−1^) for new dynamics to occur.

## Appendix B Equal decay parameters

Here we consider the case when the parameters *α*_*i*_ are not unique. For simplicity we will consider the case when they are all equal to each other, i.e. *α*_*i*_ = *α* recognizing that sub cases mixing equality and inequality will result in a combination of this analysis and that presented in Section 3. The reduced model from (6) is

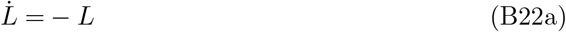

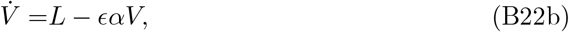

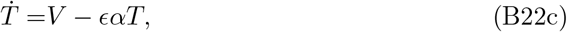

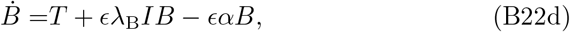

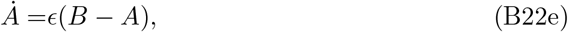

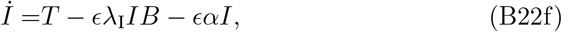

subject to initial conditions *V* (0) = *T*(0) = *B*(0) = 0, *L*(0) = 1, *A*(0) = 𝒜, and *I*(0) = ℐ. We note that we have removed the equation for *F* as it is unchanged from Sections 3.1 and 3.2 where *F* ∼ *T*. We have also removed the equation for *C* since when *α*_T_ = *α*_C_ = *α* then *C* = *T* As with (6) we can solve directly for *L, V*, and *T*,

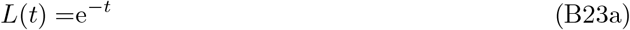

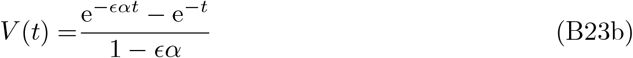

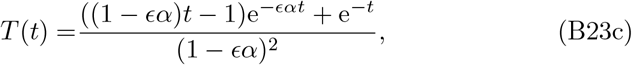

and if we knew *B* then the antibodies would still be given by (8). Thus we need only to solve for *B* and *I* in (B22) which are analyzed through each of the timescale regimes in Section 3.

We begin following Section 3.2 where *t* ∼ 𝒪(1). The original expansion of *T* when *α*_V_ ≠ *α*_T_ given by (13) is not impacted by the singularity structure of *α*_V_ = *α*_T_. As such the solution when the parameters are equal is the same as that in Section 3.2 when *α*_V_ = *α*_T_ = *α*. Since the loss of asymptotic consistency was not related to the *α*_*i*_ parameters we still have breakdown when *t* ∼ 𝒪(*ϵ*^−1*/*3^), and for the *t*_3_ timescale in Section 3.3, once again the singularity structure does not play a role when expanding *T*. Furthermore, since we only considered the leading order behaviour which did not depend on *α*_V_ or *α*_T_ then the solutions for *B* and *I* are the same to leading order, (26) and (24) respectively.

The *t*_4_ timescale of Section 3.4, where peak and decay of antibody occurs, is where the *α*_*i*_ parameters have a dominant role and the singularity structure appears in (27) for example. When the parameters are the same then substituting *t* = *ϵ*^−1^*t*_4_ into (B23c) and expanding for *ϵ* ≪ 1 yields

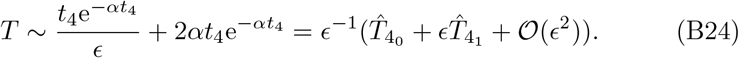

From here, the setup follows that of Section 3.4 with the equations to solve given by (28) leading to interleukin quasi-steady state (29) and differential equation for the plasma B-cells given by (30) with the change in all three that 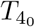 is replaced by 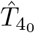. The plasma B-cell solution becomes

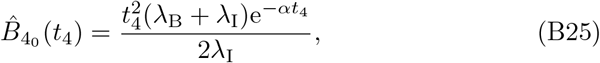

which we observe still satisfies the required (32) for *t*_4_ ≪ 1. From (33) we obtain the antibody concentration

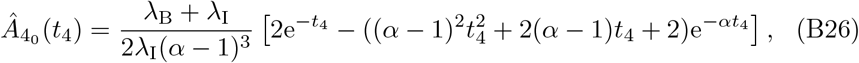

which itself has a singularity issue if *α* = 1. In this particular case the solution reduces to

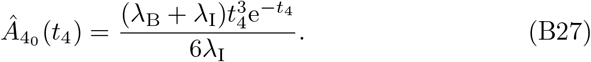

When all of the parameters are equal then min(*α*_V_, *α*_T_, *α*_B_) = min(*α*_V_, *α*_T_) = *α* and so the interleukin cessation timescale described in Section 3.5 should occur as the exponential decay rates for both CD4^+^ T-cells and plasma B-cells are identical. However, the identical decay rates also introduces algebraic terms and so the quasi-steady interleukin concentration (29) becomes

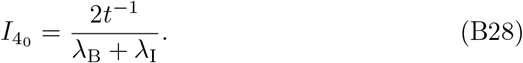

We observe the interesting result that when the parameters are equal the interleukin concentration still decays to zero, but with a power law decay rather than an exponential one.

The reduction of the solution in the *t*_4_ timescale allows an easy analysis of the maximal plasma B-cell concentration where, from (B25), we conclude that the maximal concentration of plasma B-cells occurs when *t*_4_ = 2*α*^−1^ with maximal B-cell concentration 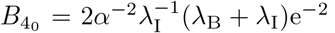. For general *α* the maximal concentration of antibody does not have a clean analytic form because of the mixture of polynomial and exponential terms in (B26). However, when *α* = 1 then the antibody concentration is given by (B27) and has a single exponential term. Upon differentiating, the maximum concentration occurs at *t*_4_ = 3 for all *λ*_B_ and *λ*_I_ with maximal concentration given by 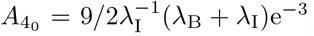. The independence of the maximal concentration time on *λ*_B_ and *λ*_I_ is discussed in Section 4.

## Notes

### Competing Interest Statement

The authors have declared no competing interest.

### Funding Statement

This study was funded by grants received from the Natural Sciences and Engineering Research Council of Canada

### Author Declarations

Data is used in this paper for model verification. The data comes from previously published research and can also be accessed at https://github.com/ionmoles/mRNA-inhost

### Summary of Updates

Revised based on feedback from peer review

